# Durability of ChAdOx1 nCov-19 (AZD1222) vaccination in people living with HIV - responses to SARS-CoV-2, variants of concern and circulating coronaviruses

**DOI:** 10.1101/2021.09.28.21264207

**Authors:** Ane Ogbe, Mathew Pace, Mustapha Bittaye, Timothy Tipoe, Sandra Adele, Jasmini Alagaratnam, Parvinder K Aley, M. Azim Ansari, Anna Bara, Samantha Broadhead, Anthony Brown, Helen Brown, Federica Cappuccini, Paola Cinardo, Wanwisa Dejnirattisai, Katie J. Ewer, Henry Fok, Pedro M. Folegatti, Jamie Fowler, Leila Godfrey, Anna L. Goodman, Bethany Jackson, Daniel Jenkin, Mathew Jones, Stephanie Longet, Rebecca Makinson, Natalie G. Marchevsky, Moncy Mathew, Andrea Mazzella, Yama F. Mujadidi, Lucia Parolini, Claire Petersen, Emma Plested, Katrina M. Pollock, Thurkka Rajeswaran, Maheshi N. Ramasamy, Sarah Rhead, Hannah Robinson, Nicola Robinson, Helen Sanders, Sonia Serrano, Helen Stockmann, Tom Tipton, Anele Waters, Panagiota Zacharopoulou, Eleanor Barnes, Susanna Dunachie, Philip Goulder, Paul Klenerman, Gavin R. Screaton, Alan Winston, Adrian V. S. Hill, Sarah C. Gilbert, Miles Carroll, Andrew J Pollard, Sarah Fidler, Julie Fox, Teresa Lambe, John Frater

## Abstract

Duration of protection from SARS-CoV-2 infection in people with HIV (PWH) following vaccination is unclear. In a sub-study of the phase 2/3 the COV002 trial (NCT04400838), 54 HIV positive male participants on antiretroviral therapy (undetectable viral loads, CD4+ T cells >350 cells/ul) received two doses of ChAdOx1 nCoV-19 (AZD1222) 4-6 weeks apart and were followed for 6 months. Responses to vaccination were determined by serology (IgG ELISA and MesoScale Discovery (MSD)), neutralisation, ACE-2 inhibition, gamma interferon ELISpot, activation-induced marker (AIM) assay and T cell proliferation. We show that 6 months after vaccination the majority of measurable immune responses were greater than pre-vaccination baseline, but with evidence of a decline in both humoral and cell mediated immunity. There was, however, no significant difference compared to a cohort of HIV-uninfected individuals vaccinated with the same regimen. Responses to the variants of concern were detectable, although were lower than wild type. Pre-existing cross-reactive T cell responses to SARS-CoV-2 spike were associated with greater post-vaccine immunity and correlated with prior exposure to beta coronaviruses. These data support the on-going policy to vaccinate PWH against SARS-CoV-2, and underpin the need for long-term monitoring of responses after vaccination.

## Introduction

The global COVID-19 pandemic has led to over 200 million cases and 4.2 million deaths (1). Vaccines which have been licensed against SARS-CoV-2 include the AstraZeneca ChAdOx1 nCov-19 (AZD1222) adenoviral vectored vaccine, of which over 1 billion doses have been made available worldwide. People living with HIV (PWH) represent a high-risk group for adverse clinical outcomes from viral infections such as influenza and COVID-19, with some evidence for higher hospitalisation and mortality rates (2–6). This can in part be attributed to a state of immune cell depletion and chronic immunopathology including immune activation and exhaustion which is only partially restored by antiretroviral therapy (ART)(7, 8). Studies on influenza and tetanus toxin vaccination in PWH have shown that antibody levels post-vaccination were dependent on CD4 T cell count and activated T follicular helper (Tfh) cell frequencies, which can vary widely in PWH (9, 10), resulting in broader concerns over reduced responses to vaccines (11) and specific vaccination guidelines for PWH (12). Some studies also report that vaccination of PWH may induce immune activation and reactivate the HIV reservoir (13, 14).

ChAdOx1 nCov-19 containing SARS-CoV-2 full length spike has been shown to induce potent humoral and cellular immune response in vaccine recipients (15–18). We recently reported the safety and immunogenicity of the ChAdOx1 nCoV-19 vaccine in PWH up to 2 months post initial vaccination (16) and the durability of T and B cell responses following natural infection with SARS-CoV-2 (19). There are, however, few studies evaluating the durability of immunity following vaccination against COVID-19 (20, 21). A recent open label phase I trial showed durable SARS-CoV-2 T and B cell immune response up to 6 months following vaccination in adults without HIV using a low dose of mRNA vaccine mRNA-1273 (21), with similar results in another study using standard mRNA-1273 dosing (22). There have been no studies to date reporting the durability of immune responses in PWH.

Since the rollout of COVID-19 vaccines, divergent mutations in the viral sequence in the original SARS-CoV-2 strain have given rise to the alpha (B.1.1.7), beta (B.1.351), gamma (P.1), and more recently delta (B.1.617.2) variants of concern (VOCs). Infections with VOCs have become dominant in several countries (23). Studies of symptomatic disease in fully vaccinated individuals report variable effectiveness (ChAdOx1 nCoV-19 – alpha (74%), delta (67%) and BNT162b2 – alpha (93.7%), delta (88%)), but with evidence for sustained protection from severe disease (24–26). Nonetheless, breakthrough infections have been recorded and a significant proportion of the world’s population remains unvaccinated (27). Understanding the ability of immune responses generated in PWH to recognise VOCs is key to informing vaccination strategies, especially in vulnerable populations.

Pre-existing cross-reactive T and B cell responses in individuals naïve to SARS-CoV-2 infection and vaccination to the circulating common cold coronaviruses (CCC) HKU1, OC43, 299E and NL63 have been identified (28–34), however the impact of this cross-reactivity is unclear. While some reports point to a beneficial role in mitigating disease severity and the induction of neutralising antibodies in both vaccination and natural infection (33, 35, 36), others report no biological function (37, 38) or a potential pathological role (39).

In this open-label, non-randomised sub-study of male participants with HIV on ART (CD4+ T cell count >350 cells/ul) receiving ChAdOx1 nCoV-19, we investigate the immunological landscape six months after vaccination. We evaluate the durability of the cellular and humoral immune response to SARS-CoV-2 and VOCs and assess the potential role of cross-reactive CCC immune responses in the modulation of post-vaccine responses, presenting evidence for an interaction with the beta coronaviruses, HKU1 and OC43.

## Results

### Participants

PWH (N=54; all male) were recruited as part of the ChAdOx1 nCoV-19 COV002 clinical trial (NCT04400838) in November 2020. Participants had undetectable VL (<50 HIV RNA copies/ml) and a median CD4 count of 694 cells/µl (IQR 573.5 – 859.5) at the time of recruitment. Most participants were of white ethnicity (81.5%). Other reported ethnicities were Asian (3.7%), mixed (7.4%) and other (7.4%). Demographically-matched HIV seronegative controls were provided from the ChAdOx1 nCoV-19 COV002 clinical trial. All participants received ChAdOx1 nCoV-19 4-6 weeks apart and were followed for 6 months **(Figure 1a, Table 1)**.

**Figure 1:**
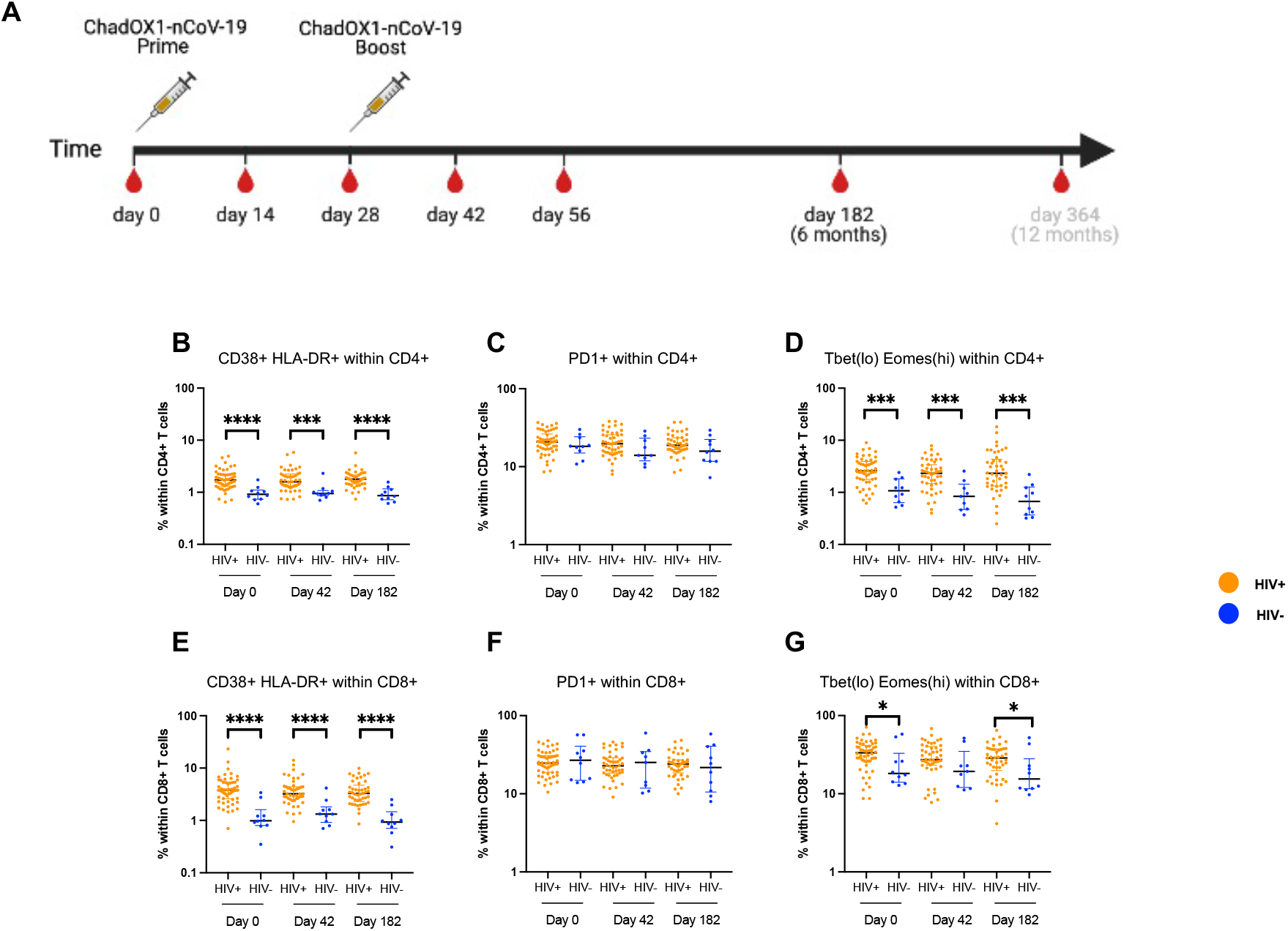
PWH show higher baseline immune activation and exhaustion. **A)** Schematic showing vaccination schedule for ChAdOx1 nCoV-19 in PWH. Frequency of **(B)** CD38+ HLA-DR+, **(C)** PD1+ **(D)** Tbet(lo) Eomes(hi) cells within CD4+ and **(E)** CD38+ HLA-DR+, **(F)** PD1+ **(G)** Tbet(lo) Eomes(hi) cells within CD8+ T cells. Comparison of two groups by two-tailed Mann-Whitney U test. Where indicated * = <0.05, ** = <0.01, *** = < 0.001 and **** = <0.0001.

**Table 1:**
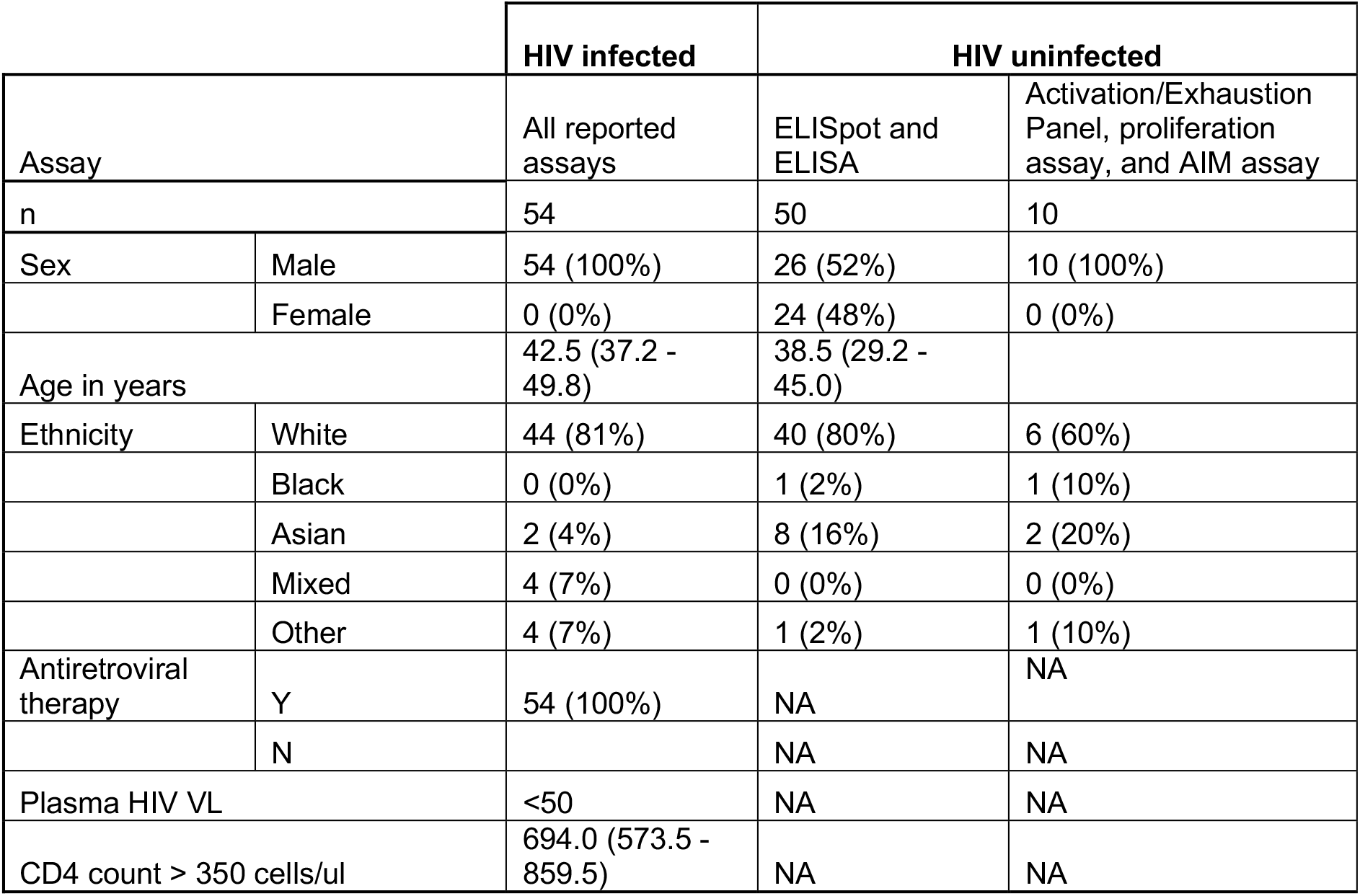
Demographic information for HIV infected and HIV uninfected receiving ChAdOx1 nCoV-19.

### Persistent immune activation in PWH before and after vaccination

T cell immune activation and exhaustion were assayed at day 0 baseline, day 42 and day 182 after first ChAdOx1 nCoV-19 vaccination **(Figure 1b-g).** There were significantly higher frequencies of CD38+ HLA-DR+ expressing CD4+ and CD8+ cells in PWH compared to HIV negative controls, consistent across all timepoints **(Figure 1b and e,** gating strategy in **supplementary figure 1a)**. There was a transient increase in the frequency of CD38+ HLA-DR+ CD4 and CD8+ T cells 14 days after vaccination in PWH which returned to pre-vaccination levels by 6 months **(Supplementary figure 1c, f)**. Expression of the immune checkpoint inhibitor PD-1 on CD4+ and CD8+ T cells was not significantly different between PWH and HIV negative controls with no statistically significant changes after vaccination **(Figure 1c and f)**. The frequency of CD4+ and CD8+ PD-1 expressing cells fluctuated early after vaccination in PWH but was restored to pre-vaccination levels at 6 months **(supplementary figure d, g, j and m).** The frequency of functionally exhausted Tbet (lo) Eomesodermin (Eomes) (hi) CD4+ and CD8+ T cells was higher in PWH compared to HIV-negative individuals both before and after vaccination. **(Figure 1d and g, supplementary figure 1e, h, k and n)**.

### Humoral immunity against ChAdOx1 nCoV-19 in PWH persists for 6 months

We previously reported detectable antibody levels up to 56 days following ChAdOx1 nCoV-19 vaccination in PWH (16). To determine the further persistence of antibody responses, total IgG for spike (S), receptor binding domain (RBD) and nucleocapsid (N), as well as neutralising antibody levels were measured at days 0 and day 182. Two independent ELISA technologies were used for binding IgG assays: a standardised in-house total IgG against spike and Meso Scale Discovery (MSD) binding assays measuring S, RBD and N antibody levels. Levels of anti-spike IgG measured using the two assays were positively correlated (**Supplementary figure 2a and b**, r=0.7, p<0.0001 and r = 0.9, p<0.0001 at Days 0 and 182, respectively; Spearman). At day 182 post-vaccination, antibodies to S and RBD but not N were significantly higher than at baseline (S: day 0 = 3/43 [6.9%], day 182 = 35/42 [83.3%]; RBD: day 0 = 0/43 [0%] and day 182 = 27/42 [64.2%]) (**Figure 2a – c**), consistent with observed responses being driven by vaccination rather than infection.

**Figure 2:**
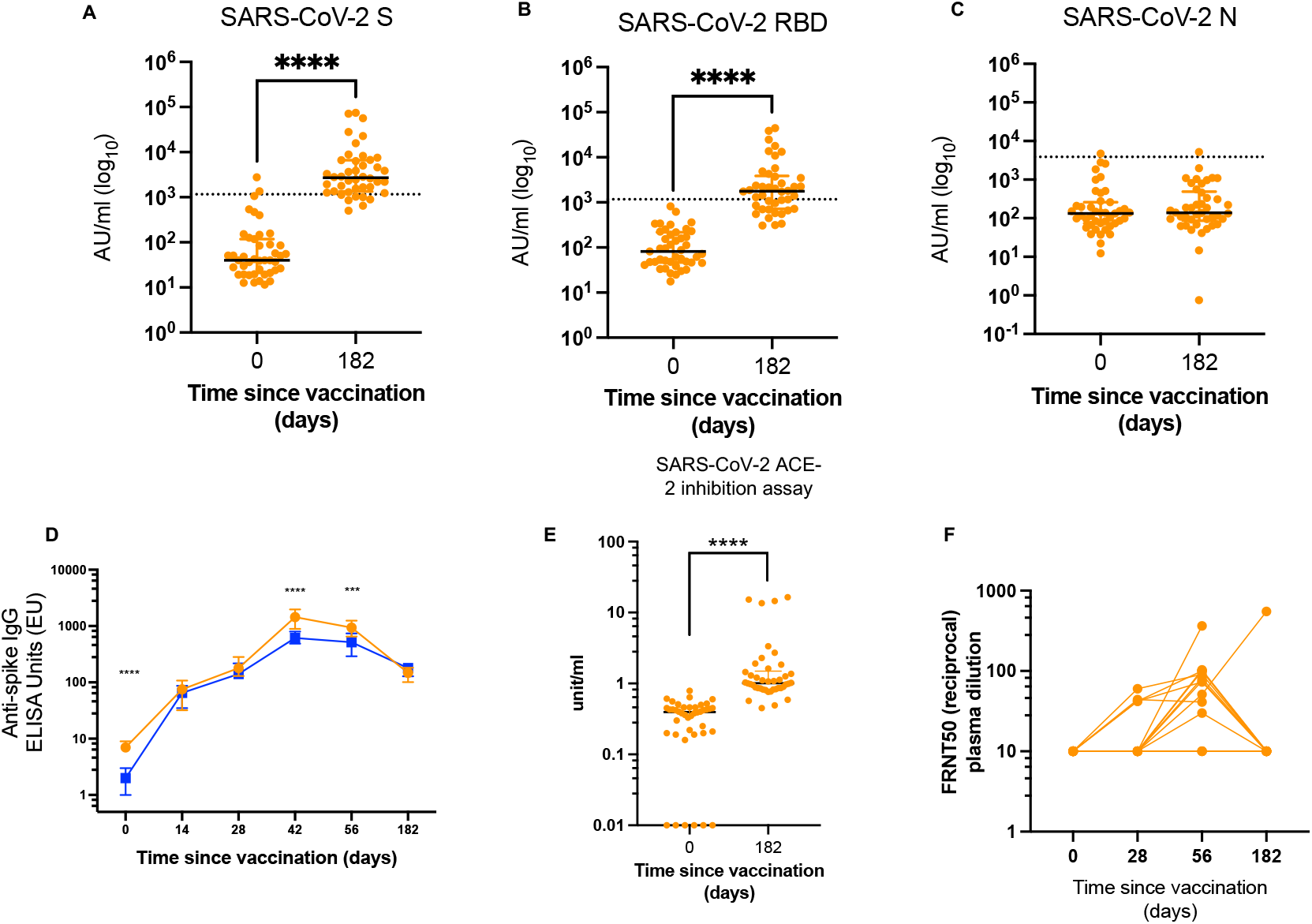
Antibody levels against SARS-CoV-2 6 months after ChAdOx1 nCoV-19 vaccination. IgG levels for SARS-CoV-2 **(A)** Spike **(B)** RBD and **(C)** N protein measured at day 0 (baseline) and day 182 (6 months post-vaccination) using the MSD ELISA assays. **(D)** Comparison between antibody kinetics in HIV+ and HIV-across all available timepoints. **(E)** ACE-2 inhibition assay at baseline and 6 months post-vaccination and **(F)** Live-virus focus reduction neutralisation assay (FRNT) on n =15 HIV+ donors on day 0, 28, 56 and 182. Comparison of two timepoints within the same group was done by Wilcoxon matched pair sign ranked test. Comparison of two groups was done by two-tailed Mann-Whitney U test. Where indicated * = <0.05, ** = <0.01, *** = < 0.001 and **** = <0.000. Dotted lines in A – C indicate cut off points determined for each SARS-CoV-2 antigen (S, RBD and N) based on pre-pandemic sera + 3X SD.

Importantly, there was no difference in anti-spike antibody titres in HIV+ and HIV-matched participants measured at 182 days after first vaccination, although with some waning of responses in both groups after Day 56 **(Figure 2d; Supplementary Figure 2d,e)**. Pre-vaccine baseline antibody titres correlated positively with early post-vaccination timepoints at day 14, and 28 but not 42, 56 and 182 (**Supplementary figure 2f, Supplementary table 1)**.

We next assessed the ability of antibodies from plasma collected 6 months after vaccination to compete with SARS-CoV-2 for binding to ACE-2 using an ACE-2 inhibition assay and to neutralise SARS-CoV-2 using a live virus focus reduction neutralisation assay (FRNT). FRNT was performed in a randomly selected subset of the cohort for whom we have previously reported neutralisation antibody levels up to day 56 (16). At day 182 post-ChAdOx1 nCoV-19 prime, antibodies capable of blocking the SARS-CoV-2 ACE-2 interaction were present at significantly higher levels than at pre-vaccination baseline **(Figure 2e)** and correlated strongly with anti-RBD antibodies **(Supplementary figure 2c).** However, at the same time point antibody neutralisation measured by FRNT live virus assay revealed titres below the assay detection limit in nearly all participants (13/14; 92%) **(Figure 2f)**.

### Durable SARS-CoV-2 specific T cell responses are induced following ChAdOx1 nCoV-19 vaccination

Durability of vaccine-induced SARS-CoV-2-specific T cell immunity at 6 months was assessed by IFNγ ELISpot and T cell proliferation assays. SARS-CoV-2 spike-specific ELISpot responses were maintained for 6 months in PWH following vaccination and were equivalent to the HIV negative control group **(Figure 3a and b; Supplementary figure 4a)**.

**Figure 3:**
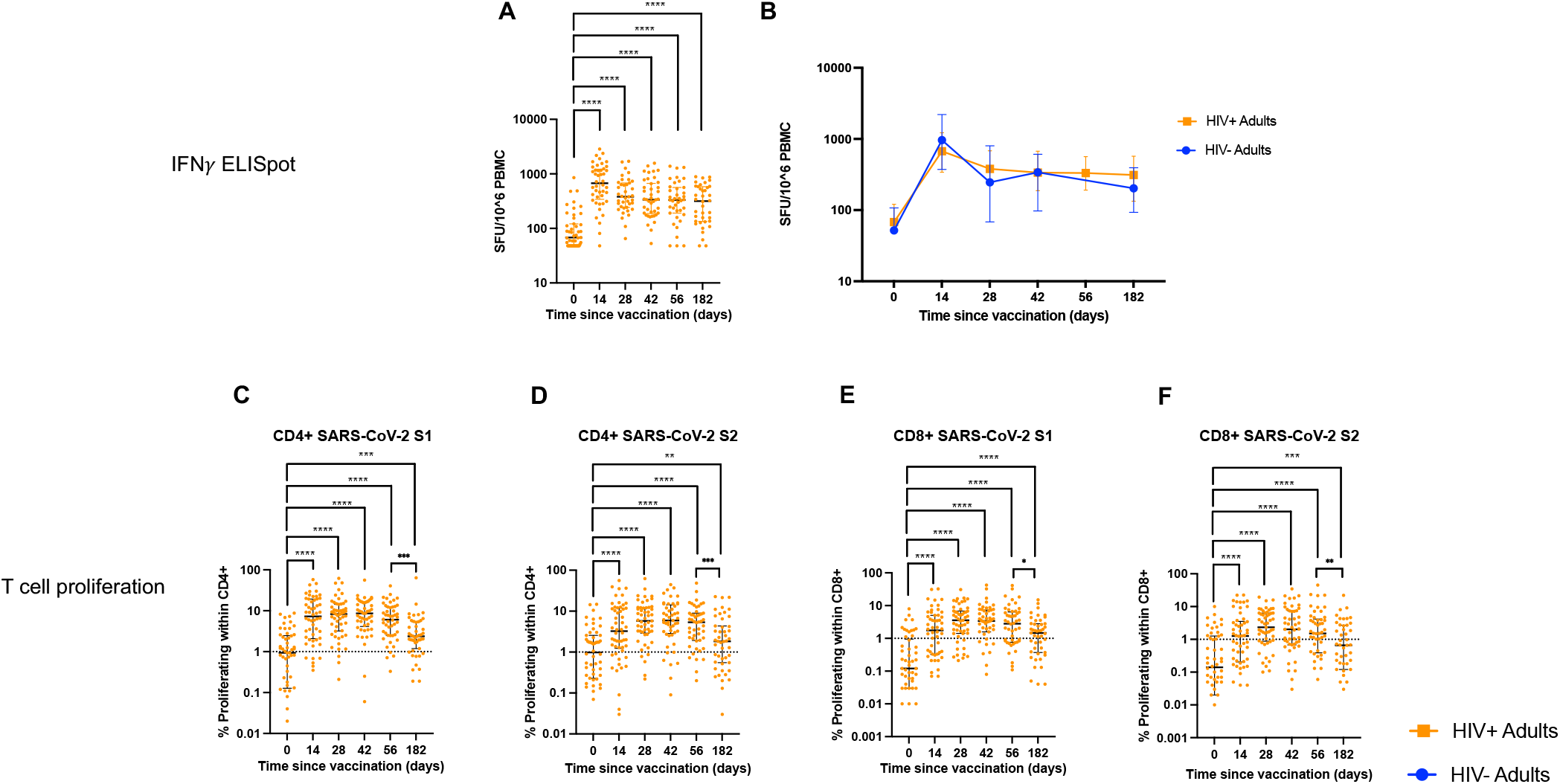
T cell responses following ChAdOx1 nCoV-19 vaccination are durable in PWH. **(A)** T cell response measured using peptides pools against SARS-CoV-2 S1 and S2 antigens by IFNγ ELISpot across all timepoints. **(B)** comparative analysis of IFNγ T cell responses in HIV+ and HIV-volunteers. Proliferative T cell responses to **(C)** SARS-CoV-2 S1 and **(D)** SARS-CoV-2 S2 in CD4+ T cells across all available timepoints. Proliferative T cell responses to **(E)** SARS-CoV-2 S1 and **(F)** SARS-CoV-2 S2 in CD8+ T cells across all available timepoints. Comparison of two timepoints within the same group was done by Wilcoxon matched pair sign ranked test. Comparison of two groups was done by two-tailed Mann-Whitney U test. Where indicated * = <0.05, ** = <0.01, *** = < 0.001 and **** = <0.000. Dotted lines in C - F indicate threshold for true positive based mean of DMSO controls + 3x SD.

For further resolution of the durability of T cell immunity, we used a T cell proliferation assay which also allows distinction of different CD4 and CD8 T cell lineage responses. The spike peptide pool was separated into S1 and S2. Gating strategy is shown in **Supplementary Figure 3a**. The frequency of SARS-CoV-2 spike-specific proliferative CD4+ and CD8+ T cell responses in PWH following vaccination were maintained at levels significantly higher than at baseline for 6 months **(Figure 3c - f)**. Longitudinal responses to FECT controls remained unchanged while PHA responses were back to baseline by day 182 **(Supplementary figure 3b and c)**. There was no difference in the magnitude of the vaccine-specific T cell proliferative responses between the HIV+ and the HIV-cohorts **(Supplementary Figure 4f - l).** Although T cell responses in PWH measured by IFNγ ELISpot peaked at day 14 and were then maintained to day 182, proliferative responses peaked later at day 42 and then contracted, such that day 182 responses were significantly lower than those measured at day 56 **(Figure 3c-f).** These kinetics are similar to those observed with the anti-S antibody response **(Supplementary figure 2d and e).**

### Vaccine-reactive T cells are not differentially biased to a specific CD4+ subset

Using CCR6 and CXCR3 expression to quantify Th1, Th2 and Th17 cells, we interrogated the phenotype of circulating T cells following vaccination (gating strategy in **Supplementary Figure 1a**). At 6 months post ChAdOx1 nCoV-19 vaccination, we found redistributions in the phenotype of the CD4+ T cells in HIV+ volunteers with increases in Th1 (CXCR3+ CCR6-) and Th2 (CXCR3-CCR6-) **(Figure 4a and b)** but not Th17 (CXCR3-CCR6+) or Tfh (CXCR5+ CD4+) **(Figure 4c and d).** None of these populations correlated with anti-spike antibody levels 6 months after infection. Although the hierarchy in cellular composition of the CD4+ T cell subsets was similar in the HIV+ and HIV-cohorts, we found circulating frequencies of Th2 subsets to be reduced while Th1 and Tfh subsets were significantly increased 6 months after vaccination in PWH **(Figure 4e, Supplementary Figure 5a** – for HIV-control data).

**Figure 4:**
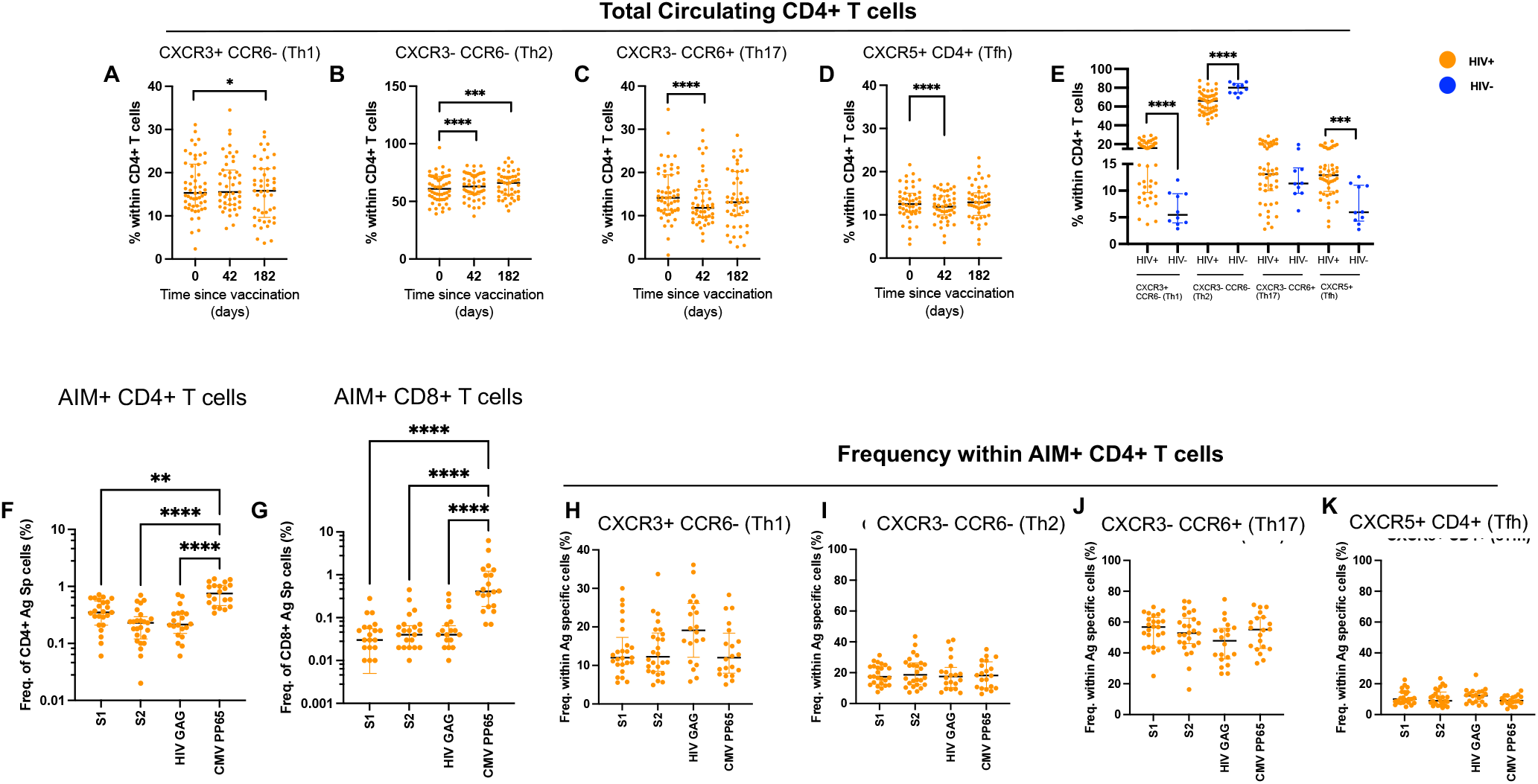
SARS-CoV-2 specific T cells are not preferentially biased for any CD4+ T cell subsets. *Ex vivo* frequencies of **(A)** CXCR3+ CCR6- (Th1), **(B)** CXCR3- CCR6- (Th2), **(C)** CXCR3- CCR6+ (Th17), and **(D)** CXCR5+ within CD4+ T cells in HIV+ volunteers measured at day 0, 42 and 182 using ex vivo T cell phenotyping. **(E)** comparative analysis of frequencies of *ex vivo* CD4+ T cell frequencies in HIV+ and HIV- volunteers at day 182 (6 months post vaccination). Measurement of frequencies of antigen specific T cells including SARS-CoV-2 S1 and S2, HIV gag and CMVpp65 using activation induced marker (AIM) assay in **(F)** CD4+ and **(G)** CD8+ T cells. For CD4 T cells, antigen specific cells were: CD25+ CD134(OX40)+ and CD25+ CD137+ and CD25+ CD69+; for CD8+ T cells, antigen specific cells were: CD25+ CD137+ and CD25+ CD69+ Frequencies of **(H)** CXCR3+ CCR6-(Th1), **(I)** CXCR3- CCR6- (Th2), **(J)** CXCR3- CCR6+ (Th17), and **(K)** CXCR5+ CD4+ T cells within antigen specific (AIM+) T cells in HIV+ volunteers. Comparison of two timepoints within the same group was done by Wilcoxon matched pair sign ranked test. Comparison of two groups was done by two-tailed Mann-Whitney U test. Where indicated * = <0.05, ** = <0.01, *** = < 0.001 and **** = <0.000.

The activation induced marker (AIM) assay was used to determine the phenotype of vaccine-specific CD4+ T cells 6 months after ChAdOx1 nCoV-19 vaccination (gating strategy in **Supplementary figure 5b**). Vaccine responses were compared with concurrent HIV Gag and CMV responses **(Figure 4f and g).** Although AIM+ cells for all antigens tested showed a Th17 bias **(Supplementary Figure 5c)**, similar to HIV-Gag or CMVpp65-specific T cells there was no preferential skewing of the SARS-CoV-2-specific T responses to any CD4 T helper subset 6 months after vaccination **(Figure 4h - k)**.

### Responses to variants of concerns (VOCs) are preserved 6 months after vaccination

Humoral and cellular immune responses to the major VOCs were measured 6 months after vaccination. Inhibition of ACE-2 binding for alpha, beta and gamma variants was increased compared to pre-vaccination baseline **(Figure 5a)**, however there was statistically significant reduction in ACE-2 inhibition for all three VOCs compared to the original SARS-CoV-2 strain, which was more apparent in the beta and gamma variants **(Figure 5b)**. T cell proliferative responses to VOCs were comparable to the SARS-CoV-2 original strain, except for SARS-CoV-2 CD4 responses to S2 which were moderately reduced across all VOCs tested **(Figure 5c – f)**. HIV+ and HIV-participants had similar magnitudes of T cell responses to S1 and S2 spike proteins of all VOCs tested, with the exception of the CD8+ SARS-CoV-2 T cell proliferative response targeting the S2 protein of the delta variant which showed a modest reduction in HIV+ participants compared to HIV-controls **(Figure 5g – j)**.

**Figure 5:**
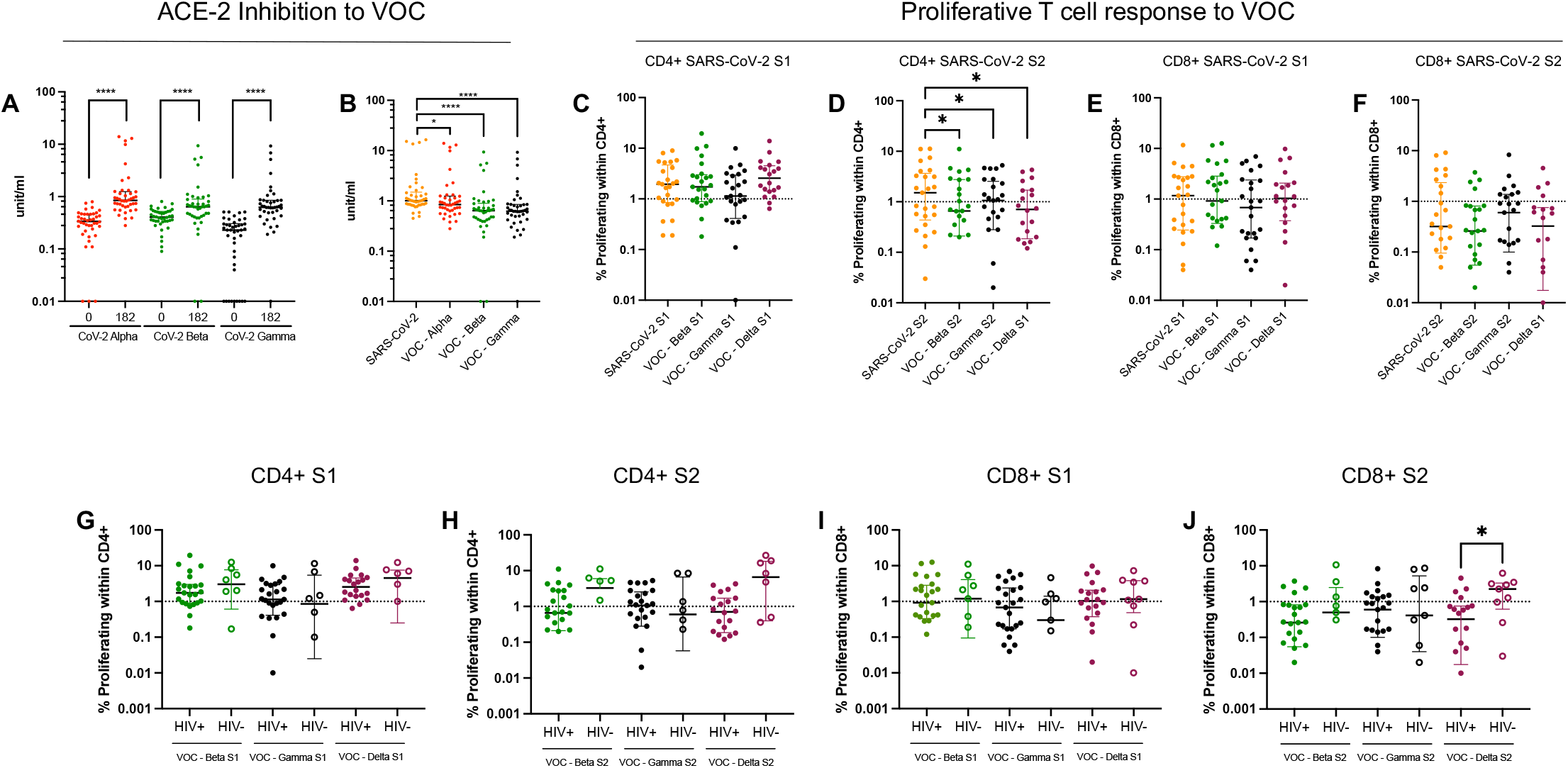
Responses to VOCs are preserved at 6 months post ChAdOx1 nCoV-19 vaccination in PWH. **(A)** ACE-2 binding inhibition assay for alpha, beta and gamma VOCs measured at day 0 (baseline) and at day 182 (6 months post-vaccination) in HIV+ volunteers. **(B)** Comparison between ACE-2 binding inhibition of SARS-CoV-2 WT strain and alpha, beta and gamma VOCs in HIV+ volunteers. Comparison between proliferative T cell responses to SARS-CoV-2 WT strain and beta, gamma and delta VOCs in **(C)** CD4+ S1, **(D)** CD4+ S2, **(E)** CD8+ S1 and **(F)** CD8+ S2 in HIV+ volunteers. Comparative analysis of **(G)** CD4+ S1, **(H)** CD4+ S2, **(I)** CD8+ S1, and **(J)** CD8+ S2 T cells responses to VOCs in HIV+ (solid circles) and HIV-(open circles). Comparison of two timepoints within the same group was done by Wilcoxon matched pair sign ranked test. Comparison of two groups was done by two-tailed Mann-Whitney U test. Where indicated * = <0.05, ** = <0.01, *** = < 0.001 and **** = <0.000. Dotted lines in C-J indicate threshold for true positive based mean of DMSO controls + 3x SD.

### Modulation of ChAdOx1 nCoV-19 post vaccination responses by pre-existing cross-reactive immunity

SARS-CoV-2 reactive T and B cells exist in unvaccinated COVID-19 naïve individuals (**Figure 3a – e, Supplementary figure 2d and e)**. To determine whether these pre-vaccine responses might reflect cross-reactivity to endemic circulating coronaviruses of the alpha (NL63 and 299E) or beta (HKU1, OC43) genera, we also measured responses to these viruses at baseline.

Based on the T cell proliferation assay, participants were divided according to those with pre-vaccine baseline SARS-CoV-2 immune responses (‘baseline responders’, (BR)) and those without pre-existing immunity (‘baseline non-responders’, (B-NR)). Regardless of any pre-existing immunity, all donors mounted an immune response following vaccination, however BR consistently showed higher magnitude CD4+ **(Figure 6a and b)** and CD8+ T cell (**Supplementary Figure 6a and b)** responses to SARS-CoV-2 S1 and S2 at most post-vaccination timepoints. Baseline SARS-CoV-2 CD4+ S2 (and to a lesser extent S1) T cell proliferation was positively correlated with subsequent post-vaccine proliferative responses targeting the same regions **(Supplementary Figure 7b and c, supplementary table 2a - d)**, which is of potential interest as S2 is associated with regions of homology to other coronaviruses. T cell and IgG responses to the endemic CCCs (HKU1 (clade 1 and 2), OC43, 299E and NL63) in HIV-infected participants remained mostly unchanged by vaccination with ChAdOx1 nCov-19 indicating that vaccination did not boost these responses (**Figure 7c, Supplementary Figure 8 and 9)** however, IgG responses to SARS-CoV-1 and MERS-CoV in PWH were higher at 6 months **(Figure 7a,b)**.

**Figure 6:**
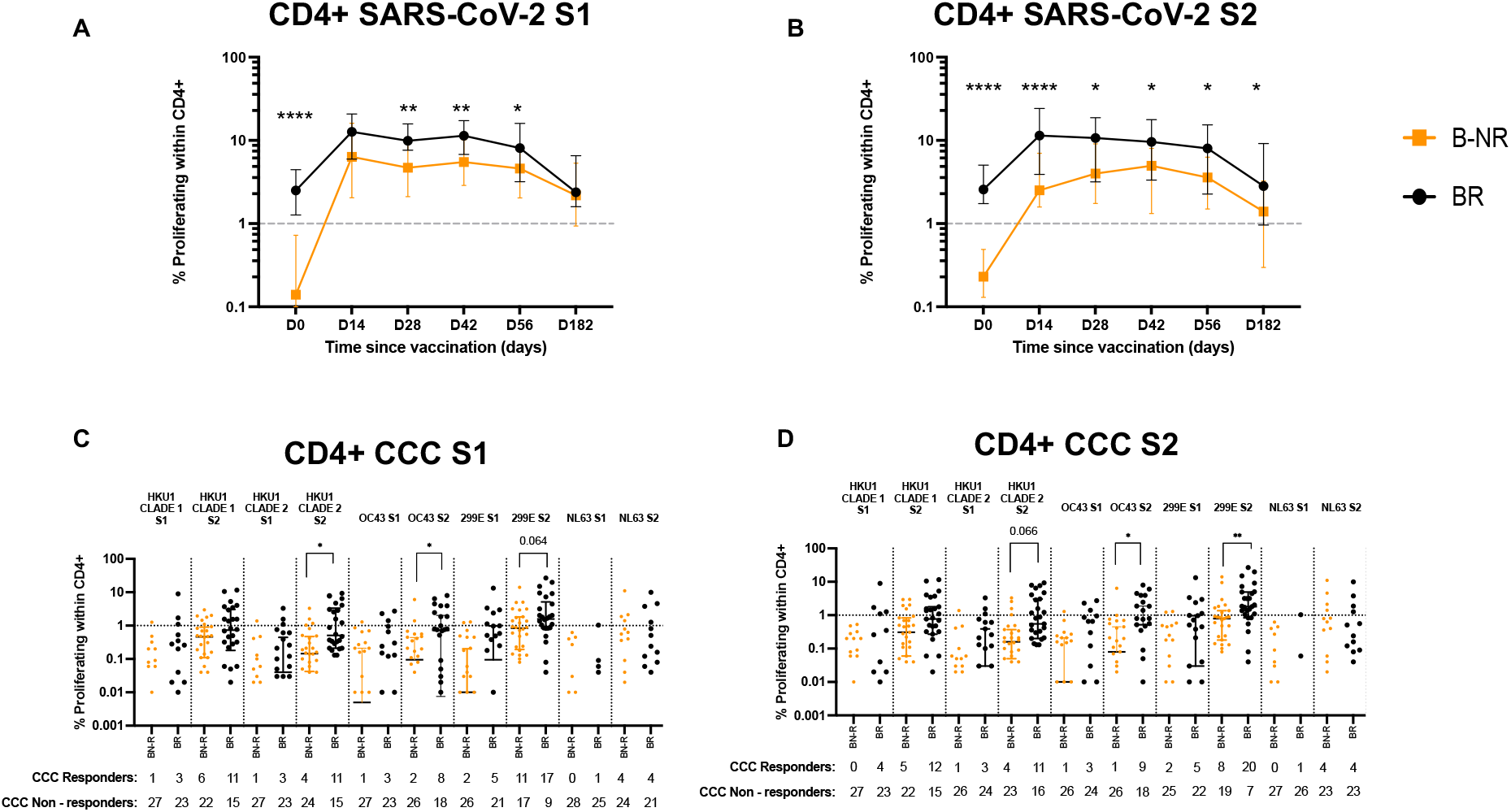
Pre-existing cross-reactive CD4+ T cell responses in PWH measured at baseline are associated with high magnitude T cell responses post ChAdOx1 nCoV-19 vaccination. Baseline CD4+ SARS-CoV-2 responses were split into baseline responders (BR, proliferation >1%, black circles and black lines) and baseline non-responders (B-NR, Proliferation <1%, yellow circles and yellow lines) and CD4 T cell responses post vaccination were analysed at all available timepoints for **(A)** SARS-CoV-2 S1 and **(B)** SARS-CoV-2 S2. T cells responses targeting **(C)** S1 and **(D)** S2 proteins in endemic CCCs are measured at baseline in BR and B-NR. Comparison of two timepoints within the same group was done by Wilcoxon matched pair sign ranked test. Comparison of two groups was done by two-tailed multiple Mann-Whitney U test. CCC responses among participants were compared using fisher’s exact test and listed in supplementary table 3. P values as indicated or * = <0.05, ** = <0.01, *** = < 0.001 and **** = <0.000. Dotted lines in indicate threshold for true positive based mean of DMSO controls + 3x SD

**Figure 7:**
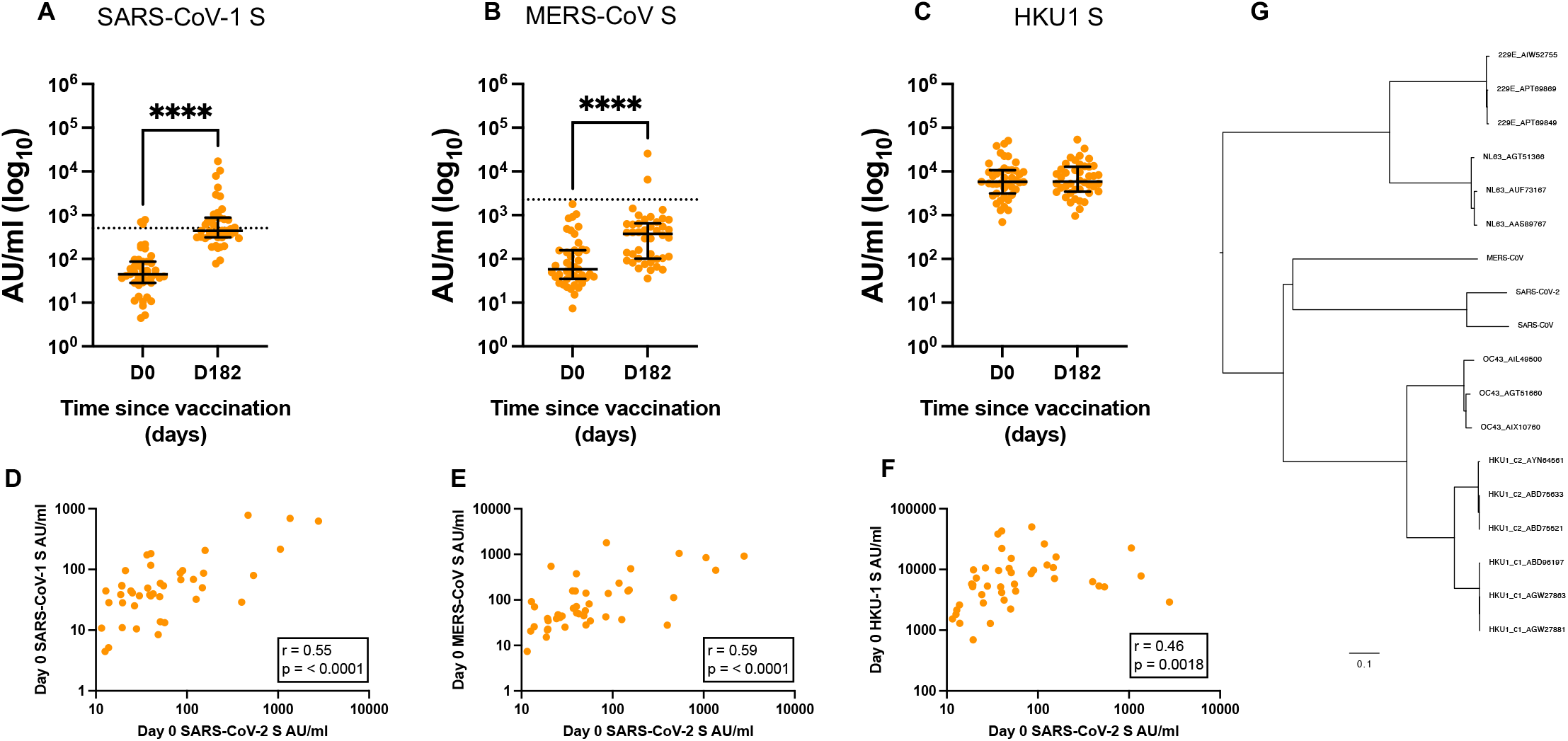
Cross reactive humoral immune responses among betaCoVs. Antibody titres against **(A)** SARS-CoV, **(B)** MERS-CoV, and **(C)** HKU-1 spike proteins measured at day 0 (baseline) and day 182 (6 months post-vaccination) in HIV+ participants. Correlation between baseline antibody titres for SARS-CoV-2 and **(D)** SARS-CoV-1, **(E)** MERS-CoV, and **(F)** HKU-1 spike protein at baseline. **(G)** Phylogenetic tree showing relationship between coronaviruses. Correlation was performed via Spearman’s rank correlation coefficient and comparison of two timepoints within the same group was done by Wilcoxon matched pair sign ranked test. Where indicated ns = not significant, * = <0.05, ** = <0.01, *** = < 0.001 and **** = <0.0001. Dotted lines in A – B indicate cut off points determined for each SARS-CoV-2 antigen based on pre-pandemic sera + 3X SD.

Focusing on baseline pre-existing responses – and dividing the cohort of PWH into the SARS-CoV-2 BR and B-NR groups as before - participants with baseline proliferative T cell responses to SARS-CoV-2 spike, also had T cell responses targeting the S2 spike regions of CCCs, especially for the beta coronaviruses HKU1 and OC43 and alpha coronavirus 299E **(Figure 6c and d, Supplementary Figure 6c and d, Supplementary table 3)**. This was supported by humoral responses taken at the same pre-vaccination timepoint, which showed strong correlations between SARS-Cov-2 spike IgG levels and those of SARS-CoV-1, MERS-CoV-1 and HKU1 (**Figure 7d-f; Supplementary Figure 9, supplementary table 4**). Phylogenetic analysis of spike sequences shows OC43 and HKU1 are the mostly closely related CCCs to SARS-CoV-2 **(Figure 7g).** These data suggest that prior exposure to beta coronaviruses and responses to the S2 homologous region may potentially be associated with larger and more persistent T cell responses following SARS-CoV-2 vaccination.

## Discussion

Long-lasting immune responses against SARS-CoV-2 will be necessary to confer protection from severe COVID-19. Although clinical management and effective antiretroviral therapy (ART) have improved long-term outcomes for PWH – especially in resource-rich countries – HIV-induced immunopathology evidenced by immune activation and exhaustion does not recover to the levels found in HIV uninfected subjects (40), raising concerns whether effective immune responses will persist after vaccination. We show here for the first time in PWH that vaccine-induced immunity to SARS-CoV-2 persists for at least 6 months by most assays, but with evidence that responses are starting to wane. There were no significant differences in responses by PWH and HIV-controls, extending the data from short-term responses reported previously (16, 41).

We confirm the persistent immune activation - and, to a lesser degree, functional exhaustion - in T cells in PWH on ART, but show that this does not impact the robust humoral and cellular immune responses to ChAdOx nCoV-19 that persist for 6 months. Reports on reactivation of the HIV reservoir and increased immune activation after vaccination in PWH are conflicting (13, 14, 42), and although we found a transient increase in the frequencies of T cells co-expressing CD38 and HLA-DR, this was restored to baseline by six months. Further studies will be needed to determine any impact on the HIV reservoir.

Vaccine design and regimen can skew the quality of the T cell response by the preferential induction of one CD4+ T helper subset over another (43–48). ChAdOx-1 nCoV-19 responses show a qualitative skew towards the Th1 phenotype, with increased IFNγ, IL-2 and TNF-producing T cells shortly after vaccination (18). Other studies in convalescent cohorts have linked a CCR6+ Th-17 cTfh phenotype with reduced disease severity (49). Similar to others (44, 50), we found antigen-specific CD4+T cells following vaccination were mostly a CCR6+ CXCR3- Th-17 phenotype. We did not find SARS-CoV-2 spike-specific CD4+ T cells biased towards any chemokine-expressing sub-population 6 months after vaccination, possibly reflecting the longer duration between vaccination and analysis than in other studies.

Understanding durability of both humoral and cellular immunity to SARS-CoV-2 – both likely key components of an effective response (49, 51, 52) - is key to understanding long-term protection. When we assessed the longevity of the humoral and cellular immunity in PWH 6 months after ChAdOx1 nCoV-19 vaccination, we found that vaccine-mediated antibodies to spike or RBD remained elevated above baseline and no different to HIV-controls. Similarly, T cell responses to spike were maintained at magnitudes above baseline and demonstrated similar kinetics to HIV- participants. Antibody function measured by ACE-2 binding inhibition was sustained at levels above pre-vaccination, however live neutralisation assays did not detect antibodies in the majority of the participants assayed at 6 months. Both assays identified the same participants as low (n = 13) and high (n = 1) responders, and the ACE-2 binding inhibition and SARS-CoV-2 RBD titres showed a strong positive correlation. We speculate that although differences in positive responses between the two functional assays could be as a result of function (neutralisation) versus antigenicity (ACE-2 binding inhibition), it could also in part, be due to assay sensitivity and differing dynamic ranges between assays.

SARS-CoV-2 convalescent plasma has been shown to have effective FC-mediated antibody functions such as antibody dependent cellular phagocytosis (ADCP), antibody dependent cellular cytotoxicity (ADCC) and complement dependent cytotoxicity (CDC) (53–55), which are more durable than neutralisation (54). Non-neutralising functions were not evaluated in this study, and therefore we cannot exclude that these are preserved in this cohort of PWH. Total spike IgG antibody and T cell proliferative responses in PWH were significantly lower at 6 months after vaccination compared to day 56. These results suggest detectable but waning T and B cell responses at 6 months. Similar findings were reported for the mRNA-1273 COVID-19 vaccine, and found to be age-dependent, pointing to immune aging as a contributing factor (20, 21). This comprehensive analysis of humoral and cellular immunity is consistent with studies of COVID-19 in healthy adults and PWH showing durable immune responses up to 7 months post infection (19, 52, 56, 57). Further follow up at 12 months and beyond will be important to determine the longer-term persistence of responses, especially when considering the value of booster doses.

The emergence of VOCs poses a potential roadblock to ending the pandemic. We found humoral immunity to VOCs at 6 months to be at titres lower than those targeting the original wild type (WT) SARS-CoV-2 strain, albeit still significantly higher than pre-vaccination levels. The magnitude of the T cell responses to VOCs were similar to those targeting the WT SARS-CoV-2 strain for most VOCs tested apart from the CD4+ S2 responses. For most of the VOCs, T cell responses in PWH did not differ from HIV- controls. Similar observations regarding humoral immunity have been made with the mRNA vaccine BNT162b2 although as most of these studies were done within 2 months of vaccination, information on durability of the response is lacking (58–60).

One study assessing T cell responses between 21 – 28 days after full BNT162b2 vaccination found no differences between WT and VOC CD4 responses (58). This study utilised a pool of spike peptide pools not parsed into its S1 and S2 regions, and only a limited panel of VOCs were analysed. Importantly, emerging data from real world effectiveness studies suggest that vaccination protects against death and severe disease, even following infection with VOCs (24, 26)

Cross-reactivity from previous CCC infection may impact the measured SARS-CoV-2 immune response after vaccination and natural infection (32, 33, 61). We identified measurable pre-vaccine antibody titres for SARS-CoV-2 S, RBD and N proteins in PWH. Pre-vaccination SARS-CoV-2 S antibody levels strongly correlated with those of contemporaneous beta coronaviruses SARS-CoV-1, MERS-CoV and HKU1 (of which only the latter is likely to have been experienced by these UK study participants), supporting the hypothesis that these titres result from previous infection with a similar coronavirus and some cross-reactivity across coronaviruses. Supporting the antibody data, the presence of cross-reactive T cells pre-vaccination (based on proliferative potential following antigen challenge) was associated with higher magnitude post-vaccination T cell responses.

There is much debate over the significance of cross-reactive responses. Studies have reported reduced disease severity in patients with CCC humoral responses and regions of high homology to CCC capable of trans-priming SARS-CoV-2 T and B cell responses (32, 33, 36). Pre-existing immunity was also shown to boost post vaccine responses in low dose mRNA-1273 vaccine (21) although an explanatory mechanism was not reported. Further investigations in large studies would be needed to fully elucidate the impact of baseline pre-existing immunity in post-vaccination response, but we find clear evidence of higher magnitude immune responses in those with cross-reactivity. Although our data suggest that responses to CCC may help augment subsequent vaccine responses against SARS-CoV-2, we have no evidence that on their own they are potent enough to impact susceptibility to COVID-19.

In summary, we present a comprehensive immunological assessment of ChAdOx1 nCoV-19 in PWH 6 months after vaccination. We show that despite persistent immune activation in PWH, PWH on ART and HIV-uninfected participants make equivalent T and B cell responses following vaccination. However, both responses showed signs of decline after 6 months. It is unknown what level of immunity is required to prevent hospitalisation and mortality, but real-world data suggest vaccination is successful in preventing severe disease and death even in the presence of transmissible and virulent VOC (24, 26). A booster dose may become necessary in the future to maintain long-term immunological memory to SARS-CoV-2 and the VOCs, especially for susceptible cohorts and we must continue to carefully monitor this going forward. Finally, we demonstrate that pre-existing SARS-CoV-2 cross-reactive immune responses to the beta coronaviruses HKU1 and to a lesser extent OC43 are associated with higher magnitude T cell responses following vaccination in PWH. Together these data continue to reinforce the policy of ensuring all PWH are offered vaccination against SARS-CoV-2.

## Methods

### Study design and cohort

The cohort studied in this analysis has been described previously (16). Briefly, the study comprised people living with HIV in an open-label non-randomised group within the larger multicentre phase 2/3 COV002 trial. The participants in this single-arm group comprised individuals with HIV who were stable on ART under routine follow-up at two London UK National Health Service (NHS) clinics and received ChAdOx1 nCoV-19 vaccination according to the schedule of attendance. Recruitment was done in HIV clinics at two centres in the UK (Imperial College NHS Trust and Guy’s and St Thomas’ NHS Foundation Trust). Inclusion criteria were age 18–55 years, a diagnosis of HIV infection, virological suppression on ART at enrolment (plasma HIV viral load <50 copies per mL), and a CD4 count of more than 350 cells/μL. The inclusion criteria for the COV002 trial have been published in full elsewhere (15). Written informed consent was obtained from all participants, and the trial was done in accordance with the principles of the Declaration of Helsinki and Good Clinical Practice. Study approval in the UK was done by the Medicines and Healthcare products Regulatory Agency (reference 21584/0424/001-0001) and the South Central Berkshire Research Ethics Committee (reference 20/SC/0145). Vaccine use was authorised by Genetically Modified Organisms Safety Committees at each participating site.

The ChAdOx1 nCoV-19 vaccine was produced as previously described (17). Participants received two standard intramuscular doses 4–6 weeks apart. For some assays and where sample availability allowed, comparison was made with age- and sex-matched participants who were HIV negative, aged 18–55 years, enrolled into the main COV002 phase 2/3 randomised clinical trial, and randomly assigned (5:1) to receive either ChAdOx1 nCoV-19 or MenACWY by intramuscular vaccination. The dose of vaccine administered was the same across both groups. Only participants receiving the ChAdOx1 nCoV-19 vaccine were used for comparison. Full details of the COV002 HIV-negative cohort have been published previously (15).

A screening visit where a full medical history, examination of all participants and blood tests to exclude biochemical or haematological abnormalities (full blood count; kidney and liver function tests) was done prior to enrolment. Participants with a history of laboratory-confirmed SARS-CoV-2 infection by anti-N protein IgG immunoassay (Abbott Architect, Abbott Park, IL, USA) at screening were excluded. For this study, visits on day 0 (vaccine prime) and 182 were the main study timepoints used for immunological analysis however for some assays other study visits - 14, 28 (vaccine boost), 42, and 56 - are presented where available. As some participants did not attend for their day 182 visit (n=6), there is a maximum of n=48 at this timepoint.

### Mesoscale Discovery (MSD) binding assays

IgG responses to SARS-CoV-2, SARS-CoV-1, MERS-CoV and seasonal coronaviruses were measured using a multiplexed MSD immunoassay. The V-PLEX COVID-19 Coronavirus Panel 3 (IgG) Kit (cat. no. K15399U) from Meso Scale Diagnostics, Rockville, MD USA. A MULTI-SPOT^®^ 96-well, 10 spot plate was coated with three SARS CoV-2 antigens (S, RBD, N), SARS-CoV-1 and MERS-CoV spike trimers, as well as spike proteins from seasonal human coronaviruses, HCoV-OC43, HCoV-HKU1, HCoV-229E and HCoV-NL63, and bovine serum albumin. Antigens were spotted at 200−400 μg/mL (MSD^®^ Coronavirus Plate 3). Multiplex MSD assays were performed as per the instructions of the manufacturer. To measure IgG antibodies, 96-well plates were blocked with MSD Blocker A for 30 minutes. Following washing with washing buffer, samples diluted 1:1,000-10,000 in diluent buffer, or MSD standard or undiluted internal MSD controls, were added to the wells. After 2-hour incubation and a washing step, detection antibody (MSD SULFO-TAG™ Anti-Human IgG Antibody, 1/200) was added. Following washing, MSD GOLD™ Read Buffer B was added and plates were read using a MESO^®^ SECTOR S 600 Reader. The standard curve was established by fitting the signals from the standard using a 4- parameter logistic model. Concentrations of samples were determined from the electrochemiluminescence signals by back-fitting to the standard curve and multiplied by the dilution factor. Concentrations are expressed in Arbitrary Units/ml (AU/ml). Cut- offs were determined for each SARS-CoV-2 antigen (S, RBD and N) based on the concentrations measured in 103 pre-pandemic sera + 3 Standard Deviations. Cut-off for S: 1160 AU/ml; cut-off for RBD: 1169 AU/ml; cut-off for N: 3874 AU/ml.

### SARS CoV-2 spike IgG ELISA

Humoral responses at baseline and following vaccination were assessed using a standardised total IgG ELISA against trimeric SARS CoV-2 spike protein as described previously^21^. In brief, ELISA plates were coated with 2 μg/mL of full-length trimerised SARS-CoV-2 spike glycoprotein and stored at 4°C overnight for at least 16 hours. After coating, plates were washed 6 times with PBS/0.05%Tween and blocked with casein for 1h at room temperature (RT). Thawed samples were treated with 10% Triton X-100 for 1h at RT and subsequently diluted in casein and plated in triplicate for incubation for 2h at RT alongside two internal positive controls (controls 1 and 2) to measure plate to plate variation. Control 1 was a dilution of convalescent plasma sample and control 2 was a research reagent for anti-SARS-CoV-2 Ab (code 20/130 supplied by National Institute for Biological Standards and Control (NIBSC)). The standard pool was used in a two-fold serial dilution to produce ten standard points that were assigned arbitrary ELISA units (EUs). Goat anti-human IgG (γ-chain specific) conjugated to alkaline phosphatase was used as secondary antibody and plates were developed by adding 4-nitrophenyl phosphate in diethanolamine substrate buffer. An ELx808 microplate reader (BioTek Instruments) was used to provide optical density measurement of the plates at 405mm. Standardised EUs were determined from a single dilution of each sample against the standard curve which was plotted using the 4-Parameter logistic model (Gen5 v3.09, BioTek). Each assay plate consisted of samples and controls plated in triplicate, with ten standard points in duplicate and four blank wells. The assay LLOQ (representing the lowest IgG titres that can be reliably and precisely quantified within a coefficient of variation of 25%) was determined mathematically. This was based on the 4-PL function of the standard curve data from 250 independent experiments and represents the EU value corresponding to the upper 95% confidence interval of the minimum asymptote of the 4-PL curve fit used for modelling the assay standard curves. The value of 13 EU was calculated as the assay LLOQ and this corresponds to an OD value of 0.2 for which the assay was demonstrated to show linearity.

### Focus reduction neutralization assay (FRNT)

Antibody neutralization was measured in a randomly selected subset of participants using a Focus Reduction Neutralization Test (FRNT), as described previously (62) where the reduction in the number of the infected foci is compared to a ‘no antibody’ negative control well. Briefly, serially diluted Ab or plasma was mixed with SARS-CoV- 2 strain Victoria and incubated for 1 hour at 37°C. The mixtures were then transferred to 96-well, cell culture-treated, flat-bottom microplate containing confluent Vero cell monolayers in duplicate and incubated for further 2 hours, followed by the addition of 1.5% semi-solid carboxymethyl cellulose (CMC) overlay medium to each well to limit virus diffusion. A focus forming assay was then performed by staining Vero cells with human anti-NP mAb (mAb206) followed by peroxidase-conjugated goat anti-human IgG (A0170; Sigma). Finally, the foci (infected cells) approximately 100 per well in the absence of antibodies, were visualized by adding TrueBlue Peroxidase Substrate. Virus-infected cell foci were counted on the classic AID ELISpot reader using AID ELISpot software. The percentage of focus reduction was calculated and IC50 (reported as FRNT50) was determined using the probit program from the SPSS package.

### MSD ACE-2 inhibition assay

A multiplexed MSD immunoassay (MSD, Rockville, MD) was used to measure the ability of human sera to inhibit ACE2 binding to SRAS-CoV-2 spike (B, B.1, B.1.1.7, B.1.351 or P.1). A MULTI-SPOT® 96-well, 10 Spot Plate (Plate 7) was coated with eight SARS-CoV-2 spike and RBD antigens (B, B.1, B.1.1.7, B.1.351 or P.1). Multiplex MSD Assays were performed as per manufacturer’s instructions. To measure ACE2 inhibition, 96-well plates were blocked with MSD blocker for 30 minutes. Plates were then washed in MSD washing buffer, and samples were diluted 1:10 and 1:100 in diluent buffer. Importantly, an ACE2 calibration curve which consists of a monoclonal antibody with equivalent activity against spike variants was used to interpolate results as arbitrary units. Furthermore, internal controls and the WHO international standard were added to each plate. After 1- hour incubation recombinant human ACE2-SULFO-TAG™ was added to all wells. After a further 1-hour plates were washed and MSD GOLD™ Read Buffer B was added, plates were then immediately read using a MESO® SECTOR S 600 Reader.

### Isolation of peripheral blood mononuclear cells (PBMC) from whole blood

PBMCs were isolated by density gradient centrifugation using Lymphoprep (Stem Cell Technologies, Cambridge, UK). Buffy coats containing PBMCs were collected and washed twice with pre-warmed R10 medium: Roswell Park Memorial Institute (also known as RPMI) 1640 medium (Sigma Aldrich, St Louis, MO, USA) supplemented with 10% heat-inactivated <u>fetal calf serum</u> (FCS; Sigma), 1 mM penicillin-streptomycin solution (Sigma), and 2 mM L-glutamine solution (Sigma). After the second centrifugation, cells were resuspended in R10 and counted using the Guava ViaCount assay (Guava Technologies Hayward, CA, USA) on the Muse Cell Analyzer (Luminex Cooperation). T-cell enzyme-linked immunospot assay (ELISpot) assays were done on freshly isolated PBMCs, and CellTrace Violet (CTV; ThermoFisher Scientific, CA, USA) T cell proliferation assay was done on cryopreserved samples.

### Ex vivo IFNg ELISpot to enumerate antigen-specific T cells

ELISpot assays were performed as described previously (17) using a validated protocol with freshly isolated peripheral blood mononuclear cells (PBMCs) to determine responses to the SARS-CoV-2 spike vaccine antigen at days 0 (before vaccination), 14, 28 (boost), 42 and 56. Assays were performed using Multiscreen IP ELISpot plates (Merck Millipore, Watford, UK) coated with 10 μg/mL human anti-IFNγ antibody and developed using SA-ALP antibody conjugate kits (Mabtech, Stockholm, Sweden) and BCIP NBT-plus chromogenic substrate (Moss Inc., Pasadena, MA, USA). PBMC were separated from whole blood with lithium heparin by density centrifugation within four hours of venepuncture. Cells were incubated for 18–20 hours in RPMI (Sigma) containing 1000 units/mL penicillin, 1 mg/mL streptomycin and 10% heat-inactivated, sterile-filtered foetal calf serum, previously screened for low reactivity (Labtech International, East Sussex, UK) with a final concentration of 10µg/ml of each peptide. A total of 253 synthetic peptides (15mers overlapping by 10 amino acids) spanning the entire vaccine insert, including the tPA leader sequence were used to stimulate PBMC (ProImmune, Oxford UK). Peptides were pooled into 12 pools for the SARS-CoV-2 spike protein containing 18 to 24 peptides, plus a single pool of 5 peptides for the tPA leader. Peptides were tested in triplicate, with 2.5 × 10^5^ PBMC added to each well of the ELISpot plate in a final volume of 100μL. Results are expressed as spot forming cells (SFC) per million PBMCs, calculated by subtracting the mean negative control response from the mean of each peptide pool response and then summing the response for the 12 peptide pools spanning S1 and S2. Staphylococcal enterotoxin B (0.02 μg/mL) and phytohaemagglutinin-L (10 μg/ mL) were pooled and used as a positive control. Plates were counted using an AID automated ELISpot counter (AID Diagnostika GmbH, algorithm C, Strassberg, Germany) using identical settings for all plates, and counts were adjusted only to remove artefacts. A lower limit of detection of 48 SFC/million PBMCs was determined based on the minimum number of spots that could be detected.

### T cell proliferation assay

T cell proliferation assay was done using cryopreserved PBMCs. Briefly, PBMCs were thawed and washed twice with 1mL of PBS followed by labelling with CTV at a final concentration of 2·5 μM for 10 min at room temperature. CTV, a DNA intercalating dye, enables the measurement of the decrease in dye concentration following each cell division in proliferating cells in response to antigenic stimulation as described previously (34). The labelling reaction was quenched with 4mL of fetal bovine serum (FBS) at 4°C and cells were resuspended in RPMI medium supplemented with 10% human blood group type AB serum (Sigma), 1 mM penicillin-streptomycin solution, and 2 mM L-glutamine solution, and subsequently plated in a 96-well round bottom plate at a plating density of 0·25 × 10^6^ cells per well in duplicate wells (total of 0·5 × 10^6^ cells per condition). Cells were stimulated with peptide pools (15nmers overlapping by 11) spanning SARS-CoV-2 spike (S1 and S2), SARS-CoV-2 variants of concern (beta, gamma and delta) and HCoVs (HKU-1 – 2 consensus clades, OC43, NL63 and 299E) at a final concentration of 1 μg/mL per peptide. For antigenic control, class 1 and 2 optimal peptides for FEC-T (flu, EBV, CMV, and tetanus) were pooled at a final concentration of 1 μg/mL per peptide. Media, containing 0·1% dimethyl sulfoxide (DMSO; Sigma) representing DMSO content in peptide pools, was used as a negative control and 2 μg/mL phytohaemagglutinin L (Sigma) was used as positive control. Cells were then incubated at 37°C, with 5% carbon dioxide and 95% humidity for 7 days, with a change of media on day 4. At the end of the incubation period, cells were stained using anti-human CD3, CD4, CD8, and a live cell discriminator (Live/Dead near Infra-red, Life Technologies; ThermoFisher Scientific, CA, USA) as in supplementary table 5. All samples were acquired using a BD Fortessa X20 (BD Bioscience, San Jose, CA, USA) or MACSQuant x10 (Miltenyi Biotec, Bergisch Gladbach, Germany). Responses above 1% were considered true positive based mean of DMSO controls + 3x SD. Specificity of the assay has been previous reported in (34). All datapoints presented represent a single participant and are presented as background subtracted data.

### AIM Assay

Cryopreserved PBMCs from 25 HIV infected subjects were used for activation induced marker (AIM) assay. Briefly, PBMCs were thawed in R10 (RPMI + 10% FCS, 1% Pen/strep and 1% L-glutamine. Cells were washed, counted, and rested for 6 hours in IMDM-10 (Iscove’s Modified Dulbecco’s Medium - Sigma, I3390 + 10% Human AB serum, 1% Pen/strep and 1% L-glutamine) and 1ul/ml of benzonase nucleases (70746-3, Merck). Following rest, cells were plated at 1-2 x10^6^ cells/well in a 96 well round bottom plate. Cells were then incubated for 24 hours at 37°C and 5% CO2. After stimulation cells were stained with the anti-human antibodies contained in **supplementary table 6**. Stained cells were fixed in 4% PFA and acquired on a BD LSR II flow cytometer. The data was analysed using FlowJo version 10 and Prism version 9. Antigen-specific CD4+ and CD8+ T cells were gated using the gating strategy described by Nielsen et al (44) and shown in **supplementary figure 5** (for CD4 T cells: CD25+ CD134(OX40)+ and CD25+ CD137+ and CD25+ CD69+; for CD8+ T cells: CD25+ CD137+ and CD25+ CD69+). Chemokine receptors CCR6 and CXCR3 were used as an unbiased way of analysing T cell skewness independent of cytokine kinetics.

### Ex vivo activation and exhaustion assays

Cryopreserved PBMCS were thawed in 30mls of RPMI media supplemented with 10% FBS, 1% Pen-strep and 1% L-glutamine (R10). Cells were counted and rested for an hour at a cell density of 2 x 10^6^ per ml of R10 in the presence of benzonase endonucleases (70746-3, Merck). Following rest, 2-3 million cells were used for each of the panels. Cells were washed in staining buffer (420201, Biolegend). This was followed by blocking FC receptors (422302, Biolegend) for 10 minutes at room temperature (RT) and live cell staining using L/D aqua (L34966, Life Technologies). All cells were then washed in preparation for antibody staining. For ex vivo immune activation panel, antibodies for assessing immune activation (as listed in supplementary table 7) was used as a cocktail and added to the cell pellet. Cells were subsequently incubated at 37°C for 15 minutes which was followed by a wash and fixation in 4% paraformaldehyde (PFA) for 10 minutes at RT. PFA was washed off and cells resuspended in PBS for acquisition on flow cytometer. For immune exhaustion panel, antibodies for surface markers were prepared in a cocktail which was added to cells and incubated for 15 minutes at 37°C (supplementary table 8). Cells were then prepared for intranuclear stain using FOXP3 fixation/permeabilisation kit (Life technologies). Briefly, 100ul of fixation buffer was added to cells and incubated at RT for 30 minutes. This was followed by cellular permeabilisation using the permeabilisation buffer contained in the aforementioned kit. Antibody cocktails were prepared in permeabilisation buffer, added to cells and allowed to incubate for 30 minutes at RT. Following staining, cells were washed and resuspended in PBS for acquisition. All data was acquired on a BD LSR II flow cytometer and fluorescent minus one (FMO) gates were used to set gates for markers of interest. Gating strategies are as shown in **supplementary figure 1a and b**

### Phylogenetic analysis

We used protein BLAST to download all human coronavirus S protein sequences from NCBI database. We then randomly chose 3 sequences for each of the human coronavirus species. HKU1 was consisted of two clades and we chose three isolates for each clade (c1 and c2). We used MAFFT to align all chosen human corona viruses, SARS-CoV, MERS-CoV and SARS-CoV-2 S protein sequences. We then calculated the pairwise distances between the sequences and built a neighbour joining tree using MATLAB.

### Statistical analysis

This study was not powered to a specific endpoint and the sample size was based on practical recruitment considerations in line with other subgroups of the COV002 study. We analysed all outcomes in all participants who received both doses of the vaccination schedule and with available samples, unless otherwise specified. We log-transformed serological, FRNT50, and ELISpot data for analysis. FRNT50 titres less than 20 were given the value 10 for statistical analysis. We present medians and IQRs for immunological endpoints. We used non-parametric analysis (Spearman’s rho) for correlations between two immunological endpoints. For comparison of two non-parametrically distributed unpaired variables, we used the Wilcoxon rank sum (Mann Whitney *U*) test. For comparison of two non-parametrically distributed paired datasets, we used the Wilcoxon matched pairs signed rank test. We used the χ^2^ test for comparison of ELISpot responses. Missing data were not imputed. We did all analyses using R (version 3.6.1 or later), and Prism 9 (GraphPad Software). The COV002 study is registered with <u>ClinicalTrials.gov</u>, <u>NCT04400838</u>, and is ongoing.

## Data Availability

not applicable

## Acknowledgements

This Article reports independent research funded by UK Research and Innovation (MC_PC_19055), Engineering and Physical Sciences Research Council (EP/R013756/1), Coalition for Epidemic Preparedness Innovations, and NIHR. We acknowledge support from Thames Valley and South Midland’s NIHR Clinical Research Network and the staff and resources of NIHR Southampton Clinical Research Facility and the NIHR Oxford Health Biomedical Research Centre. PMF received funding from the Coordenacao de Aperfeicoamento de Pessoal de Nivel Superior, Brazil (finance code 001). ALF was supported by the Chinese Academy of Medical Sciences Innovation Fund for Medical Science, China (grant number 2018- I2M-2–002). MAA is supported by the Wellcome Trust and Royal Society (220171/Z/20/Z). KJE is an NIHR Biomedical Research Centre senior research fellow. AJP and EB are NIHR senior investigators. M.C., S.L. and ToT. are funded by a U.S. Food and Drug Administration Medical Countermeasures Initiative grant 75F40120C00085. The views expressed in this publication are those of the authors and not necessarily those of the NIHR, FDA or the UK Department of Health and Social Care. We thank the volunteers who participated in this study.

## Supplementary figure legends

**Supplementary figure 1: Flow cytometry gating strategy and kinetics of cells expressing activation and exhaustion marker.**

Gating strategy for **(A)** activation panel and **(B)** exhaustion panel. All gates were set based on fluorescent minus one (FMO). Frequency of cells expressing **(C)** CD38+ HLA DR+, **(D)** PD-1+, **(E)** Eomes(hi) Tbet(lo) within CD4+ T cells on HIV+ participants and frequency of cells expressing **(F)** CD38+ HLA DR+, **(G)** PD-1+, **(H)** Eomes(hi) Tbet(lo) within CD8+ T cells on HIV+ participants. Frequency of cells expressing **(I)** CD38+ HLA DR+, **(J)** PD-1+, **(K)** Eomes(hi) Tbet(lo) within CD4+ T cells on HIV- participants and frequency of cells expressing **(L)** CD38+ HLA DR+, **(M)** PD-1+, **(N)** Eomes(hi) Tbet(lo) within CD8+ T cells on HIV- participants. Comparison of two timepoints within the same group was done by Wilcoxon matched pair sign ranked test. Where indicated * = <0.05, ** = <0.01, *** = < 0.001 and **** = <0.0001

**Supplementary figure 2: Humoral immune responses against SARS-CoV-2 in PWH.**

(A) Correlations between antibody levels measured using MSD assay and in-house total IgG ELISA at **(A)** day 0, **(B)** day 182 and **(C)** correlations between day 182 SARS-CoV-2 RBD levels and ACE-2 binding inhibition assay. **(D)** Antibody levels in HIV+ participants measured across all timepoints presented as dot plots and, **(E)** before-after plots to show individual responses. **(F)** Correlation plot showing correlation matrix between SARS-CoV-2 humoral immune response across all proteins, assays and timepoints. Correlation was performed via Spearman’s rank correlation coefficient and correlation matrix was created using corrplot package on R studio. Size and Colour of the heatmap corresponds to the correlation coefficient. Comparison of two timepoints within the same group was done by Wilcoxon matched pair sign ranked test. Where indicated * = <0.05, ** = <0.01, *** = < 0.001 and **** = <0.0001.

**Supplementary figure 3: Gating strategy and T cell response to control antigens (FECT) and mitogen (PHA).**

**(A)** Gating strategy for proliferation assay. All gates were set based on DMSO controls. Background was subtracted and responses were assigned positive if they were >1% after background subtraction. **(B)** PHA responses in CD4+ and CD8+ T cells at longitudinal timepoints. **(C)** FECT responses in CD4+ and CD8+ T cells at longitudinal timepoints. Comparison of two timepoints within the same group was done by Wilcoxon matched pair sign ranked test. Where indicated * = <0.05, ** = <0.01, *** = < 0.001 and **** = <0.000. Dotted lines in indicate threshold for true positive based mean of DMSO controls + 3x SD.

**Supplementary figure 4: Longitudinal T cell responses to SARS-CoV-2 in HIV+ and HIV- subjects following ChAdOx1 nCoV-19 vaccination.**

**(A)** IFNγ ELISpot responses in HIV- volunteers at day 0, 42 and 182. T cell proliferative response to **(B)** SARS-CoV-2 S1, **(C)** SARS-CoV-2 S2 in CD4+ T cells in HIV- volunteers and T cell proliferative response to **(D)** SARS-CoV-2 S1, **(E)** SARS-CoV-2 S2 in CD8+ T cells in HIV- volunteers. Comparison of T cell proliferative responses to **(F)** SARS-CoV-2 S1, **(G)** SARS-CoV-2 S2 in CD4+ T cells **(H)** SARS-CoV-2 S1, **(I)** SARS-CoV-2 S2 in CD8+ T cells in HIV+ and HIV negative volunteers at day 0, 42 and 182. Comparison of two timepoints within the same group was done by Wilcoxon matched pair sign ranked test. Comparison of two groups was done by two-tailed multiple Mann-Whitney U test. Where indicated * = <0.05, ** = <0.01, *** = < 0.001 and **** = <0.000. Dotted lines in indicate threshold for true positive based mean of DMSO controls + 3x SD.

**Supplementary figure 5: Phenotype of total and SARS-CoV-2 S-specific circulating CD4+ T cells**

**(A)** *Ex vivo* frequencies of **(A)** CXCR3+ CCR6+ (Th1), CXCR3- CCR6- (Th2), CXCR3- CCR6+ (Th17), and CXCR5+ within CD4+ T cells in HIV- volunteers measured at various timepoints. **(B)** gating strategy for AIM assay. All cells expressing CD25+ CD134 (OX40) and CD25+ CD137+ and CD25+ CD69+ were Boolean gated as antigen specific cells. Gating was set based on DMSO control and all responses were background subtracted. CXCR3 and CCR6 quadrant gate for antigen specific population was set based on expression in bulk CD4 T cell population. **(C)** Frequency of CD4 T cell subsets in SEB, SARS-CoV-2 S1, SARS-CoV-2 S2, HIV GAG and CMVpp65. Comparison of two timepoints within the same group was done by Wilcoxon matched pair sign ranked test. Comparison of two groups was done by two-tailed multiple Mann-Whitney U test. Where indicated * = <0.05, ** = <0.01, *** = < 0.001 and **** = <0.000.

**Supplementary figure 6: Pre-existing cross-reactive immunity in PWH measured at baseline are associated with high magnitude CD8+ T cell responses post ChAdOx1 nCoV-19 vaccination**

Baseline CD8+ SARS-CoV-2 responses were split into baseline responders (BR, proliferation >1%, black circles and black lines) and baseline non-responders (B-NR, Proliferation <1%, yellow circles and yellow lines) and CD8 T cell responses post vaccination were analysed at all available timepoints for **(A)** SARS-CoV-2 S1 and **(B)** SARS-CoV-2 S2. T cells responses targeting **(C)** S1 and **(D)** S2 proteins in endemic CCCs are measured at baseline in BR and B-NR. Comparison of two timepoints within the same group was done by Wilcoxon matched pair sign ranked test. Comparison of two groups was done by two-tailed multiple Mann-Whitney U test. CCC responses among participants were compared using fisher’s exact test and listed in supplementary table 3. P values as indicated or * = <0.05, ** = <0.01, *** = < 0.001 and **** = <0.000. Dotted lines in indicate threshold for true positive based mean of DMSO controls + 3x SD.

**Supplementary figure 7: Relationship between baseline and post vaccination. timepoints for CD4+ and CD8+ proliferative T cells.**

Correlation plots showing correlation matrix for **(A)** CD4+ SARS-CoV-2 S1, **(B)** CD4+ SARS-CoV-2 S2, **(C)** CD8+ SARS-CoV-2 S1, **(D)** CD8+ SARS-CoV-2 S2. Correlation was performed via Spearman’s rank correlation coefficient and correlation matrix was created using corrplot package on R studio. Size and Colour of the heatmap corresponds to the correlation coefficient. Where indicated * = <0.05, ** = <0.01, *** = < 0.001 and **** = <0.0001.

**Supplementary figure 8: Responses to CCC in PWH**

T cell proliferative responses to CCCs HKU-1 clade 1 and 2, OC43, 299E, NL63 S1 and S2 in CD4+ and CD8+ T cells measured on day 0, 14, 28, 42, 56 and 182. Comparison of two timepoints within the same group was done by Wilcoxon matched pair sign ranked test. P values as indicated or * = <0.05, ** = <0.01, *** = < 0.001 and **** = <0.000. Dotted lines in indicate threshold for true positive based mean of DMSO controls + 3x SD.

**Supplementary figure 9: Relationship between antibody responses for SARS-CoV-2 and CCCs spike in PWH**

**(A)** Correlation plot showing correlation matrix between different circulating HCoVs. **(B)** Antibody titres at day 0 and day 182 for OC43 Spike, 299E spike and NL63 spike proteins in HIV+ participants. Correlation was performed via Spearman’s rank correlation coefficient and correlation matrix was created using corrplot package on R studio. Size and Colour of the heatmap corresponds to the correlation coefficient. Comparison of two timepoints within the same group was done by Wilcoxon matched pair sign ranked test. Where indicated * = <0.05, ** = <0.01, *** = < 0.001 and **** = <0.0001.

## References

1. WHO. WHO Coronavirus (COVID-19) Dashboard. In: World Health Organization; 2021.

2. Mellor MM, Bast AC, Jones NR, Roberts NW, Ordóñez-Mena JM, Reith AJM, Butler CC, et al. Risk of adverse coronavirus disease 2019 outcomes for people living with HIV. AIDS (London, England) 2021;35:F1–F10.

3. Sheth AN, Althoff KN, Brooks JT. Influenza susceptibility, severity, and shedding in HIV-infected adults: a review of the literature. Clinical infectious diseases : an official publication of the Infectious Diseases Society of America 2011;52:219–227.

4. Abadom TR, Smith AD, Tempia S, Madhi SA, Cohen C, Cohen AL. Risk factors associated with hospitalisation for influenza-associated severe acute respiratory illness in South Africa: A case-population study. Vaccine 2016;34:5649–5655.

5. Tesoriero JM, Swain CE, Pierce JL, Zamboni L, Wu M, Holtgrave DR, Gonzalez CJ, et al. COVID-19 Outcomes Among Persons Living With or Without Diagnosed HIV Infection in New York State. JAMA Netw Open 2021;4:e2037069.

6. Geretti AM, Stockdale AJ, Kelly SH, Cevik M, Collins S, Waters L, Villa G, et al. Outcomes of COVID-19 related hospitalization among people with HIV in the ISARIC WHO Clinical Characterization Protocol (UK): a prospective observational study. Clin Infect Dis 2020.

7. Martin GE, Sen DR, Pace M, Robinson N, Meyerowitz J, Adland E, Thornhill JP, et al. Epigenetic Features of HIV-Induced T-Cell Exhaustion Persist Despite Early Antiretroviral Therapy. Frontiers in Immunology 2021;12:1458.

8. BHIVA. BHIVA guidelines on the use of vaccines in HIV-positive adults 2015. In. www.bhiva.org/vaccination-guidelines; 2021.

9. Kroon FP, van Dissel JT, de Jong JC, van Furth R. Antibody response to influenza, tetanus and pneumococcal vaccines in HIV-seropositive individuals in relation to the number of CD4+ lymphocytes. AIDS (London, England) 1994;8:469–476.

10. Cole ME, Saeed Z, Shaw AT, Guo Y, Höschler K, Winston A, Cooke GS, et al. Responses to Quadrivalent Influenza Vaccine Reveal Distinct Circulating CD4+CXCR5+ T Cell Subsets in Men Living with HIV. Scientific Reports 2019;9:15650.

11. Zanetti AR, Amendola A, Besana S, Boschini A, Tanzi E. Safety and immunogenicity of influenza vaccination in individuals infected with HIV. Vaccine 2002;20:B29–B32.

12. Bridges CB, Fukuda K, Cox NJ, Singleton JA. Prevention and control of influenza. Recommendations of the Advisory Committee on Immunization Practices (ACIP). MMWR Recomm Rep 2001;50:1–44.

13. Christensen-Quick A, Chaillon A, Yek C, Zanini F, Jordan P, Ignacio C, Caballero G, et al. Influenza Vaccination Can Broadly Activate the HIV Reservoir During Antiretroviral Therapy. Journal of acquired immune deficiency syndromes (1999) 2018;79:e104–e107.

14. Yek C, Gianella S, Plana M, Castro P, Scheffler K, García F, Massanella M, et al. Standard vaccines increase HIV-1 transcription during antiretroviral therapy. AIDS (London, England) 2016;30:2289–2298.

15. Ramasamy MN, Minassian AM, Ewer KJ, Flaxman AL, Folegatti PM, Owens DR, Voysey M, et al. Safety and immunogenicity of ChAdOx1 nCoV-19 vaccine administered in a prime-boost regimen in young and old adults (COV002): a single-blind, randomised, controlled, phase 2/3 trial. The Lancet 2020;396:1979–1993.

16. Frater J, Ewer KJ, Ogbe A, Pace M, Adele S, Adland E, Alagaratnam J, et al. Safety and immunogenicity of the ChAdOx1 nCoV-19 (AZD1222) vaccine against SARS-CoV-2 in HIV infection: a single-arm substudy of a phase 2/3 clinical trial. The Lancet HIV 2021.

17. Folegatti PM, Ewer KJ, Aley PK, Angus B, Becker S, Belij-Rammerstorfer S, Bellamy D, et al. Safety and immunogenicity of the ChAdOx1 nCoV-19 vaccine against SARS-CoV-2: a preliminary report of a phase 1/2, single-blind, randomised controlled trial. The Lancet 2020;396:467–478.

18. Ewer KJ, Barrett JR, Belij-Rammerstorfer S, Sharpe H, Makinson R, Morter R, Flaxman A, et al. T cell and antibody responses induced by a single dose of ChAdOx1 nCoV-19 (AZD1222) vaccine in a phase 1/2 clinical trial. Nature Medicine 2021;27:270–278.

19. Adriana T, Donal TS, Ane O, Daniel O, Connor*, Matthew P, Emily A, et al. Divergent trajectories of antiviral memory after SARS-Cov-2 infection. Research Square 2021.

20. Widge AT, Rouphael NG, Jackson LA, Anderson EJ, Roberts PC, Makhene M, Chappell JD, et al. Durability of Responses after SARS-CoV-2 mRNA-1273 Vaccination. New England Journal of Medicine 2020;384:80–82.

21. Mateus J, Dan JM, Zhang Z, Moderbacher CR, Lammers M, Goodwin B, Sette A, et al. Low dose mRNA-1273 COVID-19 vaccine generates durable T cell memory and antibodies enhanced by pre-existing crossreactive T cell memory. medRxiv 2021:2021.2006.2030.21259787.

22. Pegu A, O’Connell S, Schmidt SD, O’Dell S, Talana CA, Lai L, Albert J, et al. Durability of mRNA-1273 vaccine–induced antibodies against SARS-CoV-2 variants. Science 2021:eabj4176.

23. EpiFlu G. Tracking of Variants. In: GISAID EpiFlu; 2021.

24. Lopez Bernal J, Andrews N, Gower C, Gallagher E, Simmons R, Thelwall S, Stowe J, et al. Effectiveness of Covid-19 Vaccines against the B.1.617.2 (Delta) Variant. New England Journal of Medicine 2021.

25. England PH. SARS-CoV-2 variants of concern and variants under investigation in England Technical briefing 15. In: PUBLIC HEALTH ENGLAND UK; 2021.

26. Nasreen S, Chung H, He S, Brown KA, Gubbay JB, Buchan SA, Fell DB, et al. Effectiveness of COVID-19 vaccines against variants of concern in Ontario, Canada. medRxiv 2021:2021.2006.2028.21259420.

27. Bergwerk M, Gonen T, Lustig Y, Amit S, Lipsitch M, Cohen C, Mandelboim M, et al. Covid-19 Breakthrough Infections in Vaccinated Health Care Workers. New England Journal of Medicine 2021.

28. Mateus J, Grifoni A, Tarke A, Sidney J, Ramirez SI, Dan JM, Burger ZC, et al. Selective and cross-reactive SARS-CoV-2 T cell epitopes in unexposed humans. Science 2020;370:89.

29. Grifoni A, Weiskopf D, Ramirez SI, Mateus J, Dan JM, Moderbacher CR, Rawlings SA, et al. Targets of T Cell Responses to SARS-CoV-2 Coronavirus in Humans with COVID-19 Disease and Unexposed Individuals. Cell 2020;181:1489–1501.e1415.

30. Braun J, Loyal L, Frentsch M, Wendisch D, Georg P, Kurth F, Hippenstiel S, et al. SARS-CoV-2-reactive T cells in healthy donors and patients with COVID-19. Nature 2020;587:270–274.

31. Swadling L, Diniz MO, Schmidt NM, Amin OE, Chandran A, Shaw E, Pade C, et al. Pre-existing polymerase-specific T cells expand in abortive seronegative SARS-CoV-2 infection. medRxiv 2021:2021.2006.2026.21259239.

32. Le Bert N, Tan AT, Kunasegaran K, Tham CYL, Hafezi M, Chia A, Chng MHY, et al. SARS-CoV-2-specific T cell immunity in cases of COVID-19 and SARS, and uninfected controls. Nature 2020;584:457–462.

33. Ng KW, Faulkner N, Cornish GH, Rosa A, Harvey R, Hussain S, Ulferts R, et al. Preexisting and de novo humoral immunity to SARS-CoV-2 in humans. Science 2020;370:1339.

34. Ogbe A, Kronsteiner B, Skelly DT, Pace M, Brown A, Adland E, Adair K, et al. T cell assays differentiate clinical and subclinical SARS-CoV-2 infections from cross-reactive antiviral responses. Nature Communications 2021;12:2055.

35. Loyal L, Braun J, Henze L, Kruse B, Dingeldey M, Reimer U, Kern F, et al. Cross-reactive CD4+ T cells enhance SARS-CoV-2 immune responses upon infection and vaccination. medRxiv 2021:2021.2004.2001.21252379.

36. Shrock E, Fujimura E, Kula T, Timms RT, Lee IH, Leng Y, Robinson ML, et al. Viral epitope profiling of COVID-19 patients reveals cross-reactivity and correlates of severity. Science 2020;370:eabd4250.

37. Poston D, Weisblum Y, Wise H, Templeton K, Jenks S, Hatziioannou T, Bieniasz P. Absence of SARS-CoV-2 neutralizing activity in pre-pandemic sera from individuals with recent seasonal coronavirus infection. Clin Infect Dis 2020.

38. Anderson EM, Goodwin EC, Verma A, Arevalo CP, Bolton MJ, Weirick ME, Gouma S, et al. Seasonal human coronavirus antibodies are boosted upon SARS-CoV-2 infection but not associated with protection. Cell 2021;184:1858–1864.e1810.

39. McNaughton AL, Paton RS, Edmans M, Youngs J, Wellens J, Phalora P, Fyfe A, et al. Fatal COVID-19 outcomes are associated with an antibody response targeting epitopes shared with endemic coronaviruses. medRxiv 2021:2021.2005.2004.21256571.

40. Hileman CO, Funderburg NT. Inflammation, Immune Activation, and Antiretroviral Therapy in HIV. Current HIV/AIDS Reports 2017;14:93–100.

41. Madhi SA, Koen AL, Izu A, Fairlie L, Cutland CL, Baillie V, Padayachee SD, et al. Safety and immunogenicity of the ChAdOx1 nCoV-19 (AZD1222) vaccine against SARS-CoV-2 in people living with and without HIV in South Africa: an interim analysis of a randomised, double-blind, placebo-controlled, phase 1B/2A trial. The Lancet HIV 2021;8:e568–e580.

42. Kunisaki KM, Janoff EN. Influenza in immunosuppressed populations: a review of infection frequency, morbidity, mortality, and vaccine responses. The Lancet Infectious Diseases 2009;9:493–504.

43. Golding B, Scott DE. Vaccine strategies: targeting helper T cell responses.

44. Nielsen CM, Ogbe A, Pedroza-Pacheco I, Doeleman SE, Chen Y, Silk SE, Barrett JR, et al. Protein/AS01B vaccination elicits stronger, more Th2-skewed antigen-specific human T follicular helper cell responses than heterologous viral vectors. Cell Reports Medicine 2021;2:100207.

45. Taylor JM, Ziman ME, Canfield DR, Vajdy M, Solnick JV. Effects of a Th1- versus a Th2- biased immune response in protection against Helicobacter pylori challenge in mice. Microbial pathogenesis 2008;44:20–27.

46. Bowyer G, Grobbelaar A, Rampling T, Venkatraman N, Morelle D, Ballou RW, Hill AVS, et al. CXCR3+ T Follicular Helper Cells Induced by Co-Administration of RTS,S/AS01B and Viral-Vectored Vaccines Are Associated With Reduced Immunogenicity and Efficacy Against Malaria. Frontiers in Immunology 2018;9.

47. Bentebibel S-E, Khurana S, Schmitt N, Kurup P, Mueller C, Obermoser G, Palucka AK, et al. ICOS+PD-1+CXCR3+ T follicular helper cells contribute to the generation of high-avidity antibodies following influenza vaccination. Scientific Reports 2016;6:26494.

48. Rodda LB, Netland J, Shehata L, Pruner KB, Morawski PA, Thouvenel CD, Takehara KK, et al. Functional SARS-CoV-2-Specific Immune Memory Persists after Mild COVID-19. Cell 2021;184:169–183.e117.

49. Rydyznski Moderbacher C, Ramirez SI, Dan JM, Grifoni A, Hastie KM, Weiskopf D, Belanger S, et al. Antigen-Specific Adaptive Immunity to SARS-CoV-2 in Acute COVID-19 and Associations with Age and Disease Severity. Cell 2020;183:996–1012.e1019.

50. Juno JA, Tan H-X, Lee WS, Reynaldi A, Kelly HG, Wragg K, Esterbauer R, et al. Humoral and circulating follicular helper T cell responses in recovered patients with COVID- 19. Nature Medicine 2020;26:1428–1434.

51. Zhou R, To KK-W, Wong Y-C, Liu L, Zhou B, Li X, Huang H, et al. Acute SARS-CoV-2 Infection Impairs Dendritic Cell and T Cell Responses. Immunity 2020;53:864–877.e865.

52. Dan JM, Mateus J, Kato Y, Hastie KM, Yu ED, Faliti CE, Grifoni A, et al. Immunological memory to SARS-CoV-2 assessed for up to 8 months after infection. Science 2021;371:eabf4063

53. Tso FY, Lidenge SJ, Poppe LK, Peña PB, Privatt SR, Bennett SJ, Ngowi JR, et al. Presence of antibody-dependent cellular cytotoxicity (ADCC) against SARS-CoV-2 in COVID- 19 plasma. PLOS ONE 2021;16:e0247640.

54. Lee WS, Selva KJ, Davis SK, Wines BD, Reynaldi A, Esterbauer R, Kelly HG, et al. Decay of Fc-dependent antibody functions after mild to moderate COVID-19. Cell Reports Medicine 2021;2:100296.

55. Chen X, Rostad CA, Anderson LJ, Sun H-Y, Lapp SA, Stephens K, Hussaini L, et al. The development and kinetics of functional antibody-dependent cell-mediated cytotoxicity (ADCC) to SARS-CoV-2 spike protein. Virology 2021;559:1–9.

56. Alrubayyi A, Gea-Mallorquí E, Touizer E, Hameiri-Bowen D, Kopycinski J, Charlton B, Fisher-Pearson N, et al. Characterization of humoral and SARS-CoV-2 specific T cell responses in people living with HIV. bioRxiv 2021:2021.2002.2015.431215.

57. Wajnberg A, Amanat F, Firpo A, Altman DR, Bailey MJ, Mansour M, McMahon M, et al. Robust neutralizing antibodies to SARS-CoV-2 infection persist for months. Science 2020;370:1227.

58. Geers D, Shamier MC, Bogers S, den Hartog G, Gommers L, Nieuwkoop NN, Schmitz KS, et al. SARS-CoV-2 variants of concern partially escape humoral but not T cell responses in COVID-19 convalescent donors and vaccine recipients. Science Immunology 2021;6:eabj1750.

59. Liu Y, Liu J, Xia H, Zhang X, Fontes-Garfias CR, Swanson KA, Cai H, et al. Neutralizing Activity of BNT162b2-Elicited Serum. New England Journal of Medicine 2021;384:1466–1468.

60. Wang P, Nair MS, Liu L, Iketani S, Luo Y, Guo Y, Wang M, et al. Antibody resistance of SARS-CoV-2 variants B.1.351 and B.1.1.7. Nature 2021;593:130–135.

61. Shrock E, Fujimura E, Kula T, Timms RT, Lee IH, Leng Y, Robinson ML, et al. Viral epitope profiling of COVID-19 patients reveals cross-reactivity and correlates of severity. Science 2020;370:eabd4250.

62. Supasa P, Zhou D, Dejnirattisai W, Liu C, Mentzer AJ, Ginn HM, Zhao Y, et al. Reduced neutralization of SARS-CoV-2 B.1.1.7 variant by convalescent and vaccine sera. Cell 2021;184:2201–2211.e2207.

