## Supplementary figures for "Durability of ChAdOx1 nCov-19 (AZD1222) vaccination in people living with HIV - responses to SARS-CoV-2, variants of concern and circulating coronaviruses"

**Supplementary figure 1:** Flow cytometry gating strategy and kinetics of cells expressing activation and exhaustion markers

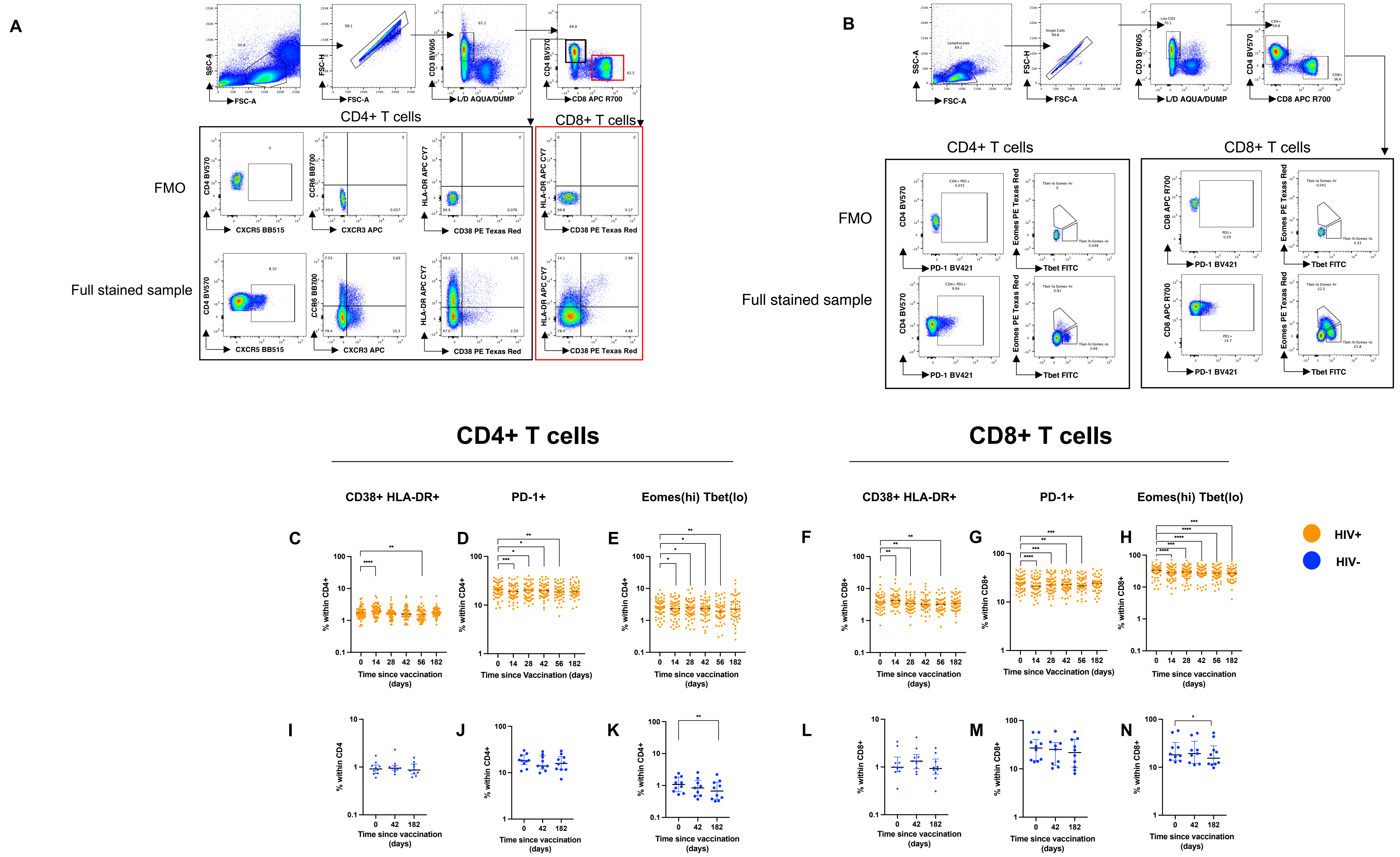

**Supplementary figure 2:** Humoral immune responses against SARS-CoV-2 in PWH.

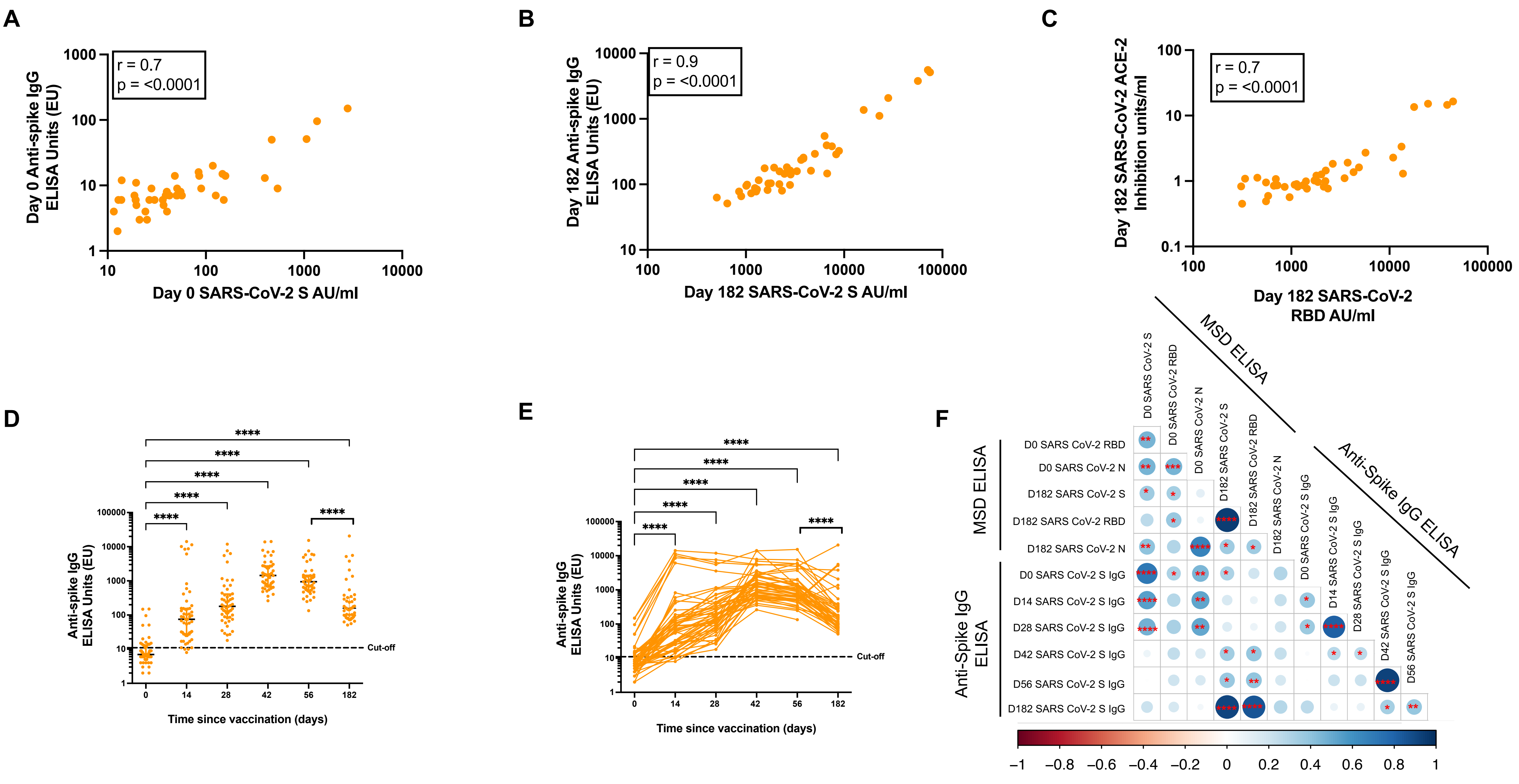

**Supplementary figure 3:** Gating strategy and T cell response to control antigens (FECT) and mitogen (PHA).

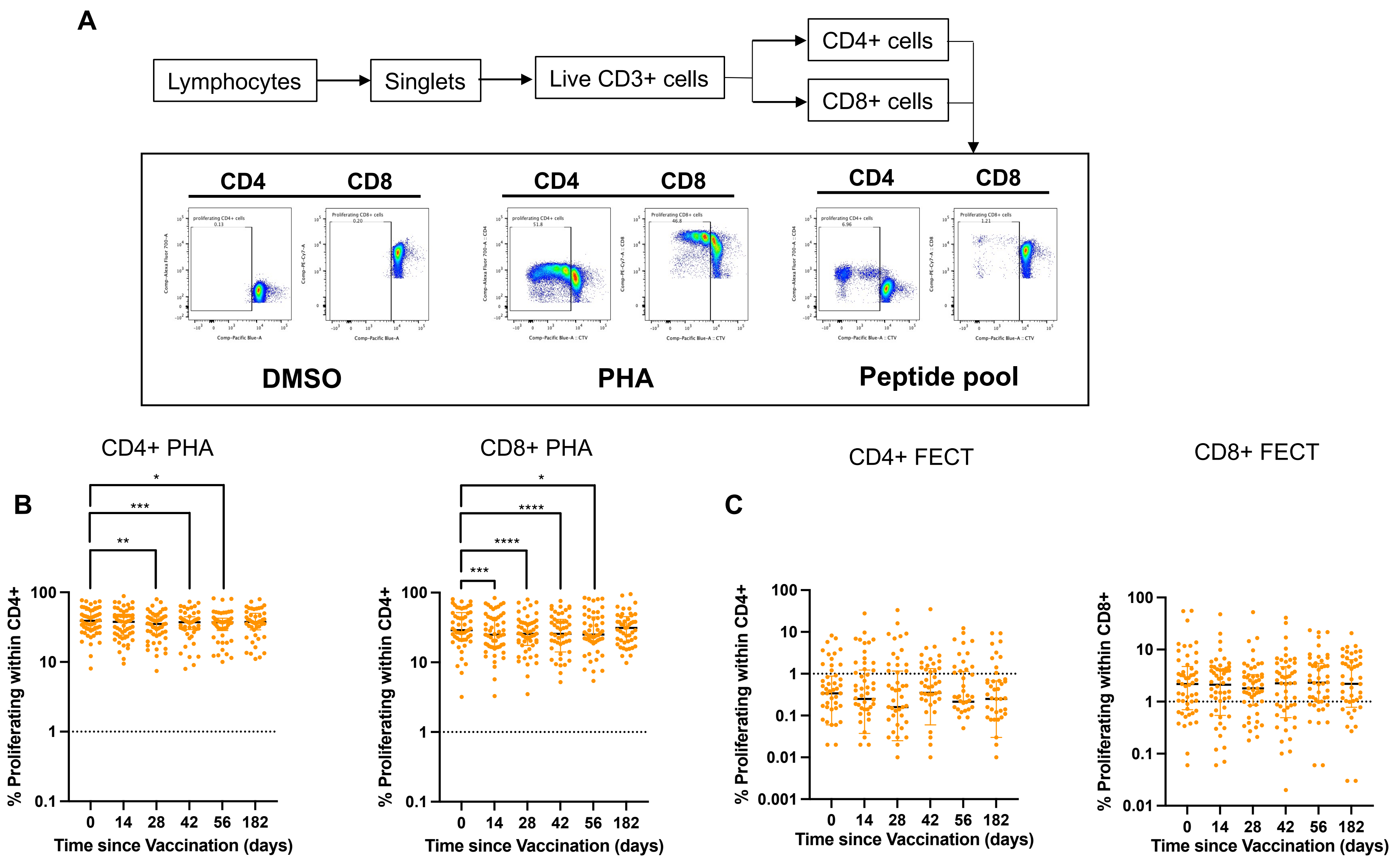

**Supplementary figure 4:** Longitudinal T cell responses to SARS-CoV-2 in HIV+ and HIV- subjects following ChAdOx1 nCoV-19 vaccination

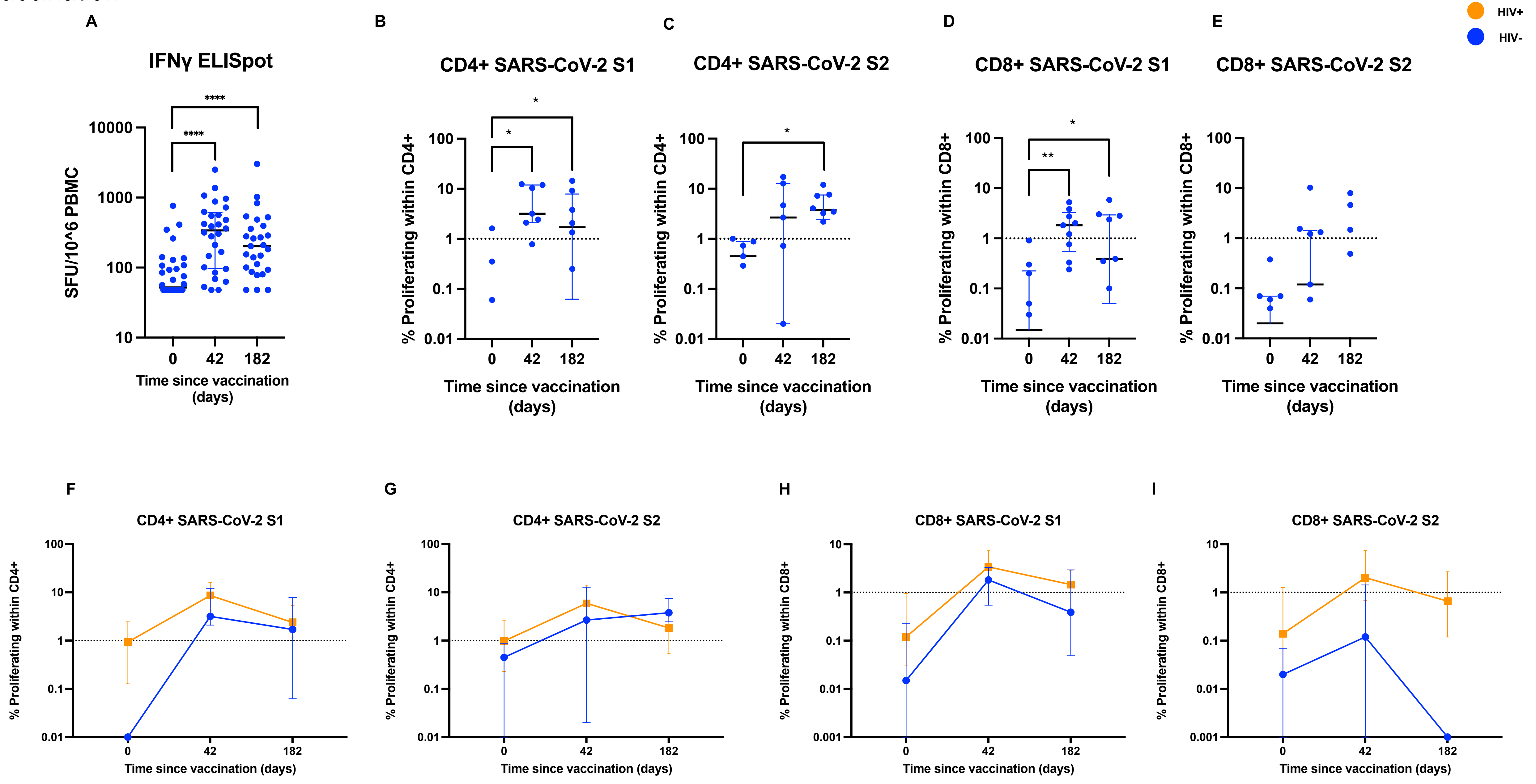

**Supplementary figure 5**: Phenotype of total and SARS-CoV-2 S-specific circulating CD4+ T cells

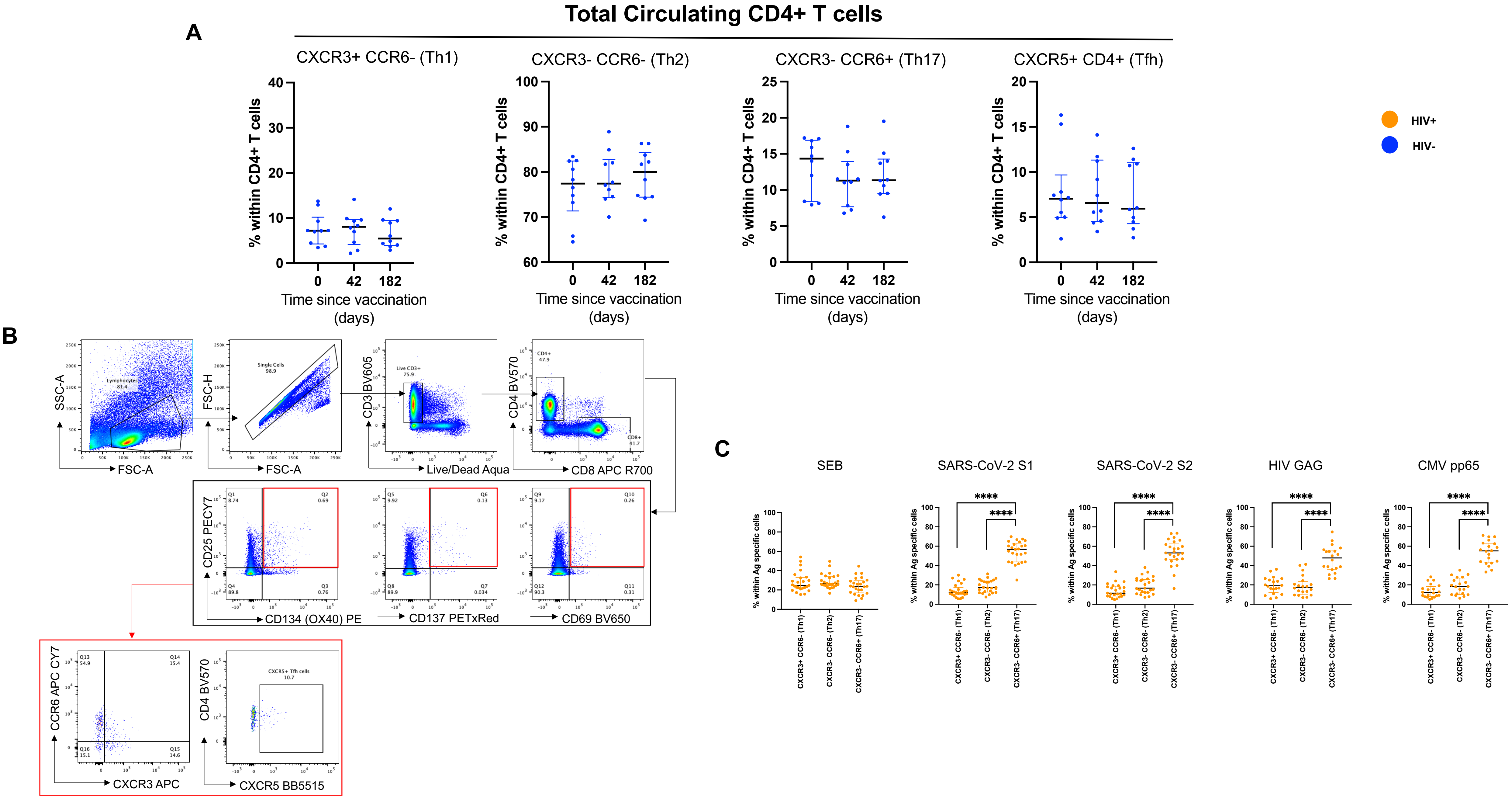

**Supplementary figure 6** : Pre-existing cross-reactive CD8+ T cell responses in PWH measured at baseline are associated with high magnitude CD8+ T cell responses post ChAdOx1 nCoV-19 vaccination

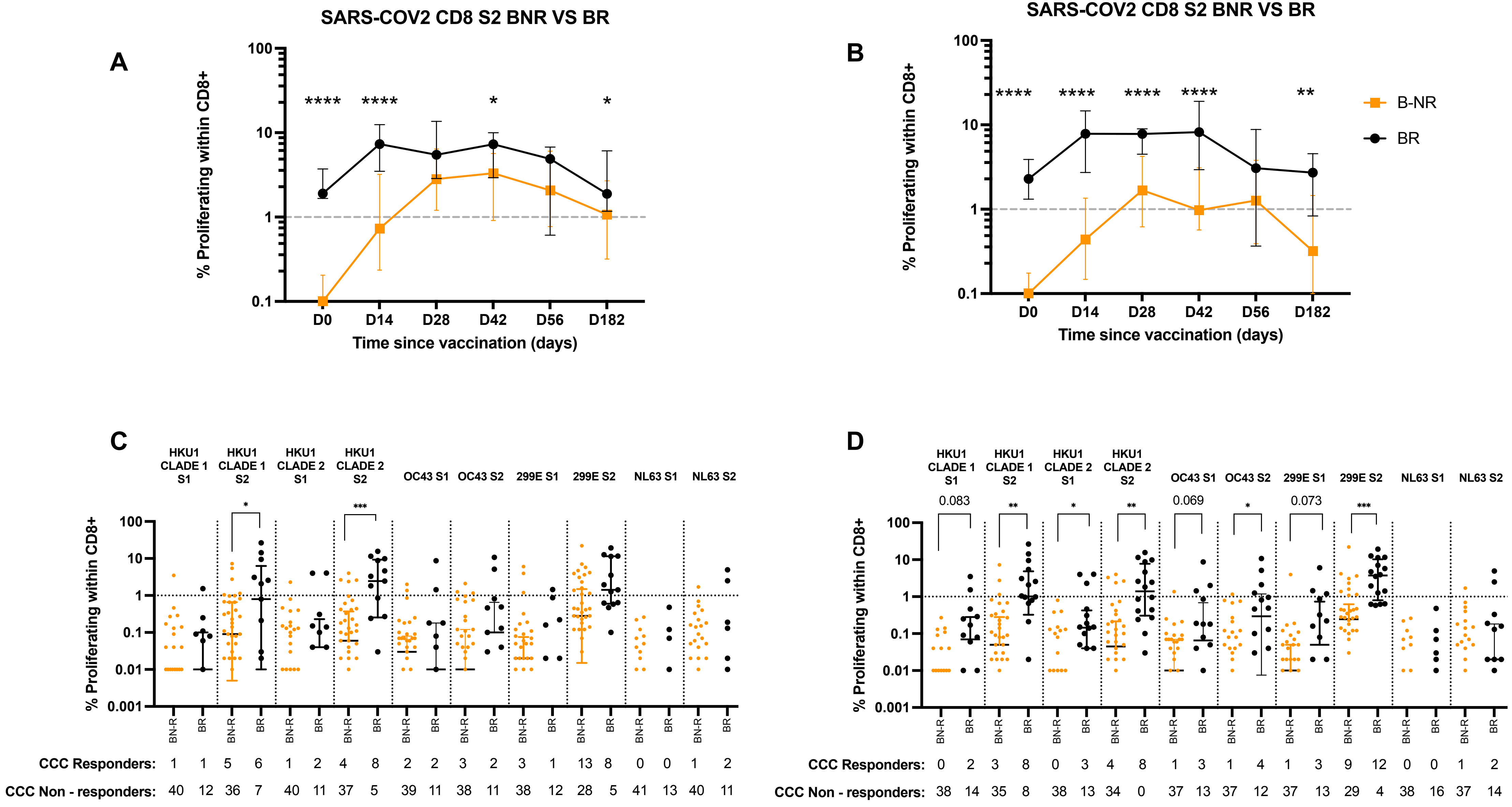

**Supplementary figure 7**: Relationship between baseline and post vaccination. timepoints for CD4+ and CD8+ proliferative T cells

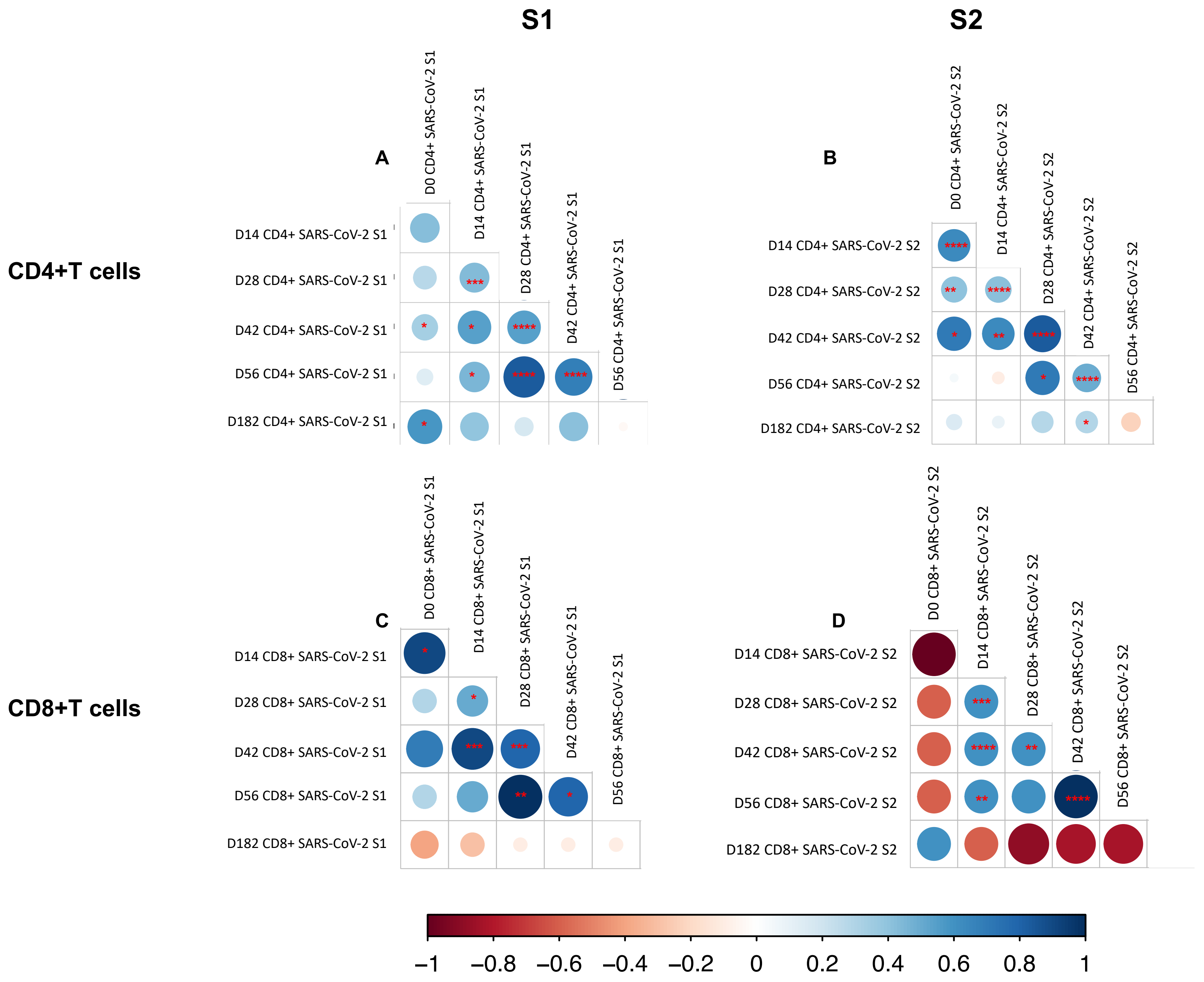

**Supplementary figure 8:** Responses to CCC in PWH

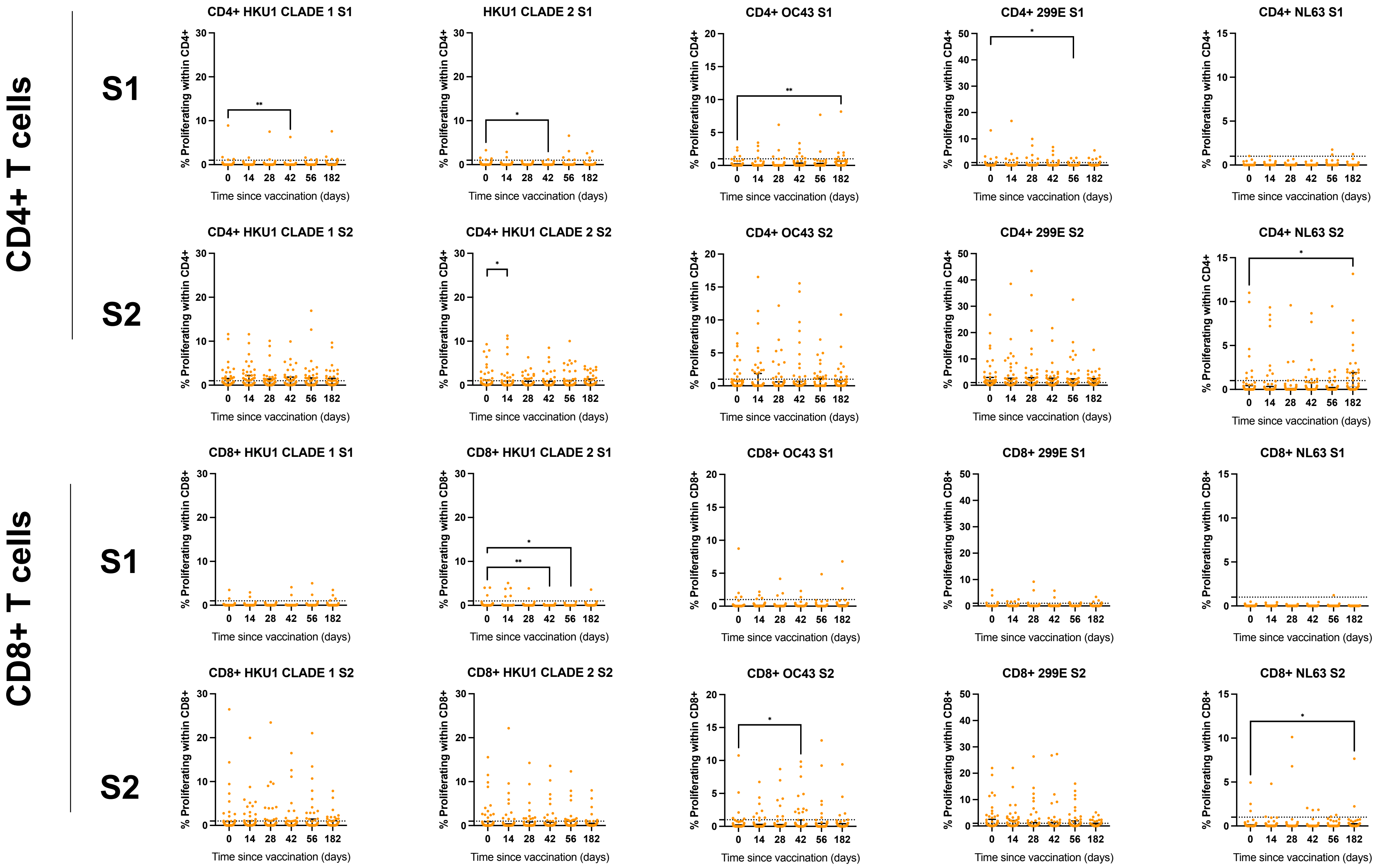

**Supplementary figure 9**: Relationship between antibody responses for SARS-CoV-2 and CCCs spike in PWH

A

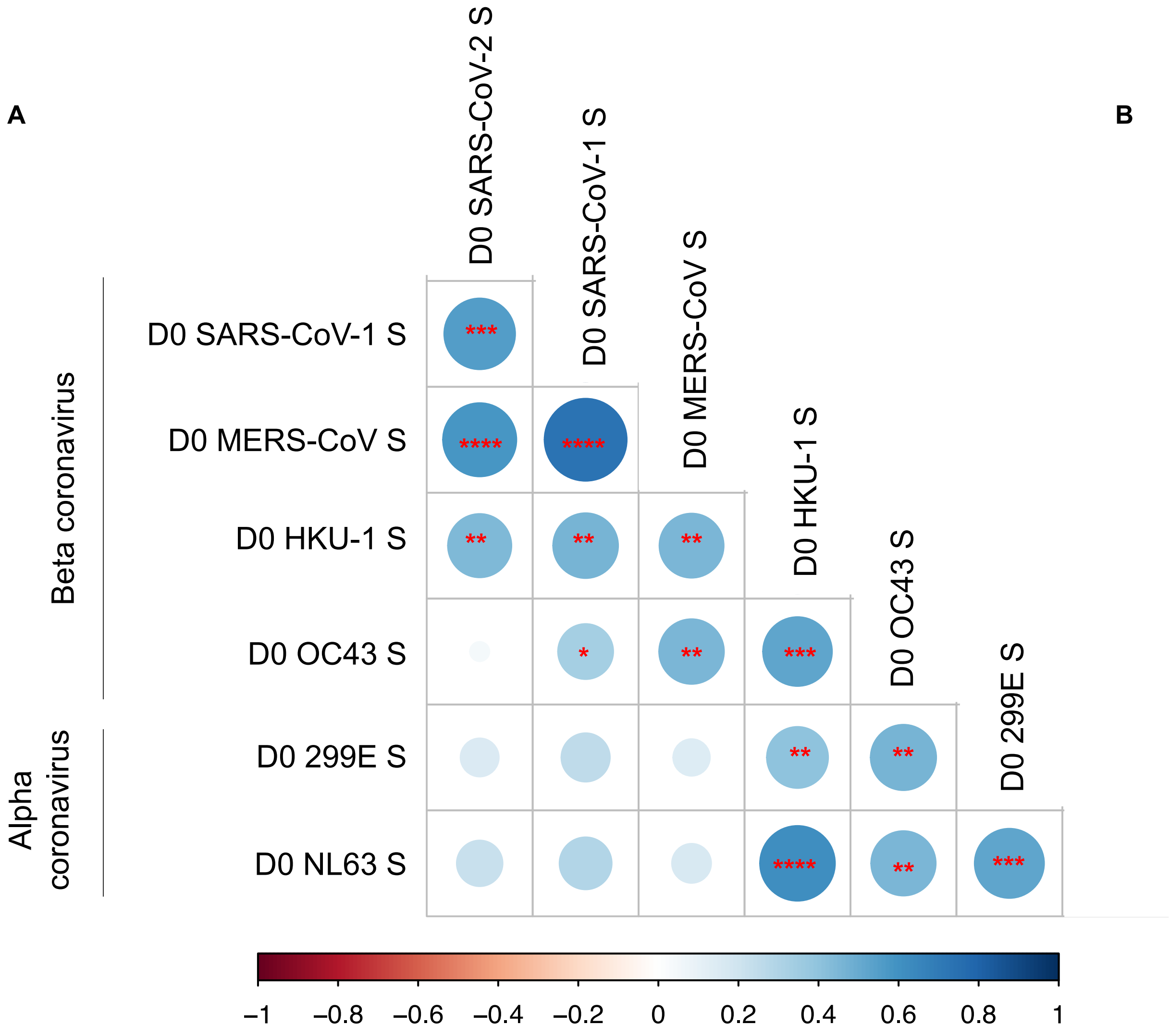


B

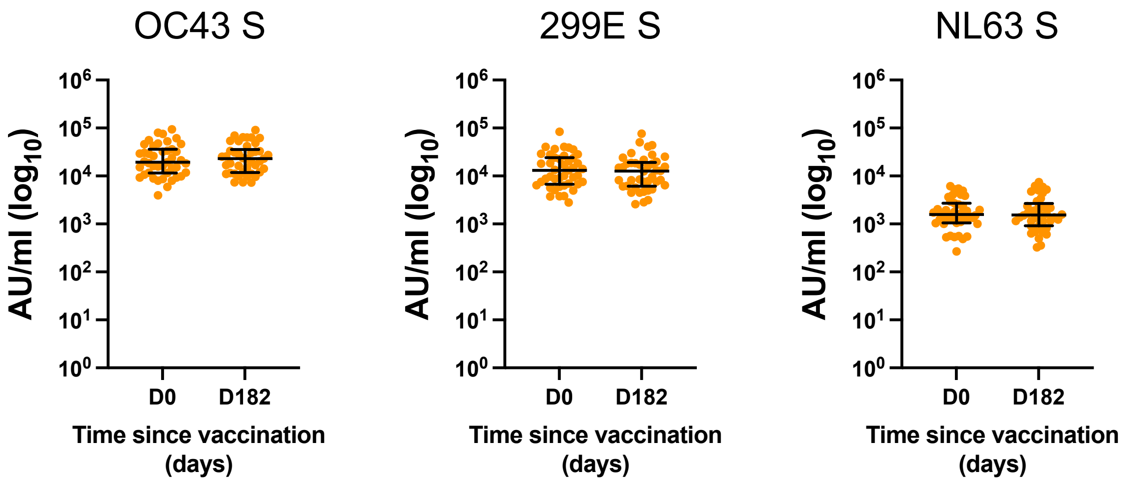
