## Supplementary tables for "Durability of ChAdOx1 nCov-19 (AZD1222) vaccination in people living with HIV - responses to SARS-CoV-2, variants of concern and circulating coronaviruses"

**Supplementary table 1:** Spearman's correlations and P values for SARS-CoV-2 specific antibody responses measured using MSD technology and anti-spike IgG.

| | | MSD_D0_SARS-CoV-2 S | MSD_D0_SARS-CoV-2 RBD | MSD_D0_SARS-CoV-2 N | MSD_D182_SARS-CoV-2 S | MSD_D182_SARS-CoV-2 RBD | MSD_D182_SARS-CoV-2 N | anti-spike IgG D0_SARS-CoV2 S IgG | anti-spike IgG D14_SARS-CoV2 S IgG | anti-spike IgG D28_SARS-CoV2 S IgG | anti-spike IgG D42_SARS-CoV2 S IgG | anti-spike IgG D56_SARS-CoV2 S IgG | anti-spike IgG D182_SARS-CoV2 S IgG |
|---|---|---|---|---|---|---|---|---|---|---|---|---|---|
| Spearman's correlation | MSD_D0_SARS-CoV-2 S | 1 | | | | | | | | | | | |
| | MSD_D0_SARS-CoV-2 RBD | 0.420433 | 1 | | | | | | | | | | |
| | MSD_D0_SARS-CoV-2 N | 0.409726 | 0.490278 | 1 | | | | | | | | | |
| | MSD_D182_SARS-CoV-2 S | 0.357117 | 0.330848 | 0.112394 | 1 | | | | | | | | |
| | MSD_D182_SARS-CoV-2 RBD | 0.262469 | 0.377846 | 0.071634 | 0.947978 | 1 | | | | | | | |
| | MSD_D182_SARS-CoV-2 N | 0.404068 | 0.292301 | 0.669679 | 0.352836 | 0.350729 | 1 | | | | | | |
| | anti-spike IgG D0_SARS-CoV2 S IgG | 0.70605 | 0.313104 | 0.443993 | 0.317133 | 0.190068 | 0.296292 | 1 | | | | | |
| | anti-spike IgG D14_SARS-CoV2 S IgG | 0.564124 | 0.223708 | 0.451171 | 0.169889 | 0.090861 | 0.232805 | 0.369578 | 1 | | | | |
| | anti-spike IgG D28_SARS-CoV2 S IgG | 0.495318 | 0.268454 | 0.457446 | 0.147413 | 0.085903 | 0.204482 | 0.359264 | 0.82714 | 1 | | | |
| | anti-spike IgG D42_SARS-CoV2 S IgG | 0.126326 | 0.188765 | 0.114547 | 0.34592 | 0.384329 | 0.234198 | 0.028872 | 0.32748 | 0.30319 | 1 | | |
| | anti-spike IgG D56_SARS-CoV2 S IgG | 0.059277 | 0.153962 | 0.116061 | 0.376079 | 0.399092 | 0.178938 | 0.001634 | 0.265899 | 0.261139 | 0.928833 | 1 | |
| | anti-spike IgG D182_SARS-CoV2 S IgG | 0.253341 | 0.119228 | 0.011176 | 0.9122 | 0.856205 | 0.28583 | 0.261829 | 0.150604 | 0.153602 | 0.382301 | 0.420977 | 1 |
| P value | MSD_D0_SARS-CoV-2 S | NA | | | | | | | | | | | |
| | MSD_D0_SARS-CoV-2 RBD | 0.004999 | NA | | | | | | | | | | |
| | MSD_D0_SARS-CoV-2 N | 0.006362 | 0.000846 | NA | | | | | | | | | |
| | MSD_D182_SARS-CoV-2 S | 0.02025 | 0.032346 | 0.478532 | NA | | | | | | | | |
| | MSD_D182_SARS-CoV-2 RBD | 0.093104 | 0.013623 | 0.652125 | 0 | NA | | | | | | | |
| | MSD_D182_SARS-CoV-2 N | 0.007958 | 0.060324 | 1.24E-06 | 0.021912 | 0.02277 | NA | | | | | | |
| | anti-spike IgG D0_SARS-CoV2 S IgG | 1.23E-07 | 0.040916 | 0.00286 | 0.040718 | 0.227958 | 0.056745 | NA | | | | | |
| | anti-spike IgG D14_SARS-CoV2 S IgG | 8.16E-05 | 0.149282 | 0.002393 | 0.282097 | 0.567144 | 0.137891 | 0.014723 | NA | | | | |
| | anti-spike IgG D28_SARS-CoV2 S IgG | 0.000733 | 0.081753 | 0.002042 | 0.351535 | 0.588566 | 0.193952 | 0.01798 | 8.13E-12 | NA | | | |
| | anti-spike IgG D42_SARS-CoV2 S IgG | 0.419547 | 0.225408 | 0.464527 | 0.024836 | 0.011974 | 0.135484 | 0.854184 | 0.032061 | 0.048114 | NA | | |
| | anti-spike IgG D56_SARS-CoV2 S IgG | 0.705742 | 0.32426 | 0.458603 | 0.014105 | 0.008841 | 0.256865 | 0.991705 | 0.084814 | 0.090754 | 0 | NA | |
| | anti-spike IgG D182_SARS-CoV2 S IgG | 0.101175 | 0.446349 | 0.943297 | 0 | 4.89E-13 | 0.066509 | 0.089873 | 0.335051 | 0.325406 | 0.011408 | 0.004937 | NA |

**Supplementary table 2a:** Spearman's correlations and P values for proliferative CD4+ T cell response to SARS-CoV-2 S1

| | | D0 CD4+ SARS-CoV-2 S1 | D14 CD4+ SARS-CoV-2 S1 | D28 CD4+ SARS-CoV-2 S1 | D42 CD4+ SARS-CoV-2 S1 | D56 CD4+ SARS-CoV-2 S1 | D182 CD4+ SARS-CoV-2 S1 |
|---|---|---|---|---|---|---|---|
| Spearman's correlation | D0 CD4+ SARS-CoV-2 S1 | 1 | | | | | |
| | D14 CD4+ SARS-CoV-2 S1 | 0.37153846 | 1 | | | | |
| | D28 CD4+ SARS-CoV-2 S1 | 0.2756917 | 0.54032014 | 1 | | | |
| | D42 CD4+ SARS-CoV-2 S1 | 0.4839074 | 0.36236934 | 0.59410277 | 1 | | |
| | D56 CD4+ SARS-CoV-2 S1 | 0.16753247 | 0.39080675 | 0.58717349 | 0.70557491 | 1 | |
| | D182 CD4+ SARS-CoV-2 S1 | 0.5252838 | 0.20527859 | 0.16472895 | 0.2553094 | 0.24133033 | 1 |
| P value | D0 CD4+ SARS-CoV-2 S1 | NA | | | | | |
| | D14 CD4+ SARS-CoV-2 S1 | 0.06744806 | NA | | | | |
| | D28 CD4+ SARS-CoV-2 S1 | 0.20291642 | 0.00018398 | NA | | | |
| | D42 CD4+ SARS-CoV-2 S1 | 0.02249512 | 0.01989648 | 2.67E-05 | NA | | |
| | D56 CD4+ SARS-CoV-2 S1 | 0.46791574 | 0.01265445 | 2.24E-05 | 2.58E-07 | NA | |
| | D182 CD4+ SARS-CoV-2 S1 | 0.02518359 | 0.25970654 | 0.34433312 | 0.14503634 | 0.17606223 | NA |

**Supplementary table 2b:** Spearman's correlations and P values for proliferative CD4+ T cell response to SARS-CoV-2 S2

| | | D0 CD4+ SARS-CoV-2 S2 | D14 CD4+ SARS-CoV-2 S2 | D28 CD4+ SARS-CoV-2 S2 | D42 CD4+ SARS-CoV-2 S2 | D56 CD4+ SARS-CoV-2 S2 | D182 CD4+ SARS-CoV-2 S2 |
|---|---|---|---|---|---|---|---|
| Spearman's correlation | D0 CD4+ SARS-CoV-2 S2 | 1 | | | | | |
| | D14 CD4+ SARS-CoV-2 S2 | 0.75395257 | 1 | | | | |
| | D28 CD4+ SARS-CoV-2 S2 | 0.58170583 | 0.6504653 | 1 | | | |
| | D42 CD4+ SARS-CoV-2 S2 | 0.51918751 | 0.47326203 | 0.6075037 | 1 | | |
| | D56 CD4+ SARS-CoV-2 S2 | 0.05912932 | 0.04398827 | 0.34522377 | 0.51920341 | 1 | |
| | D182 CD4+ SARS-CoV-2 S2 | 0.23413772 | 0.18083004 | 0.30708181 | 0.38522589 | 0.04945055 | 1 |
| P value | D0 CD4+ SARS-CoV-2 S2 | NA | | | | | |
| | D14 CD4+ SARS-CoV-2 S2 | 3.26E-05 | NA | | | | |
| | D28 CD4+ SARS-CoV-2 S2 | 0.00359574 | 1.31E-05 | NA | | | |
| | D42 CD4+ SARS-CoV-2 S2 | 0.01898251 | 0.00540799 | 8.52E-05 | NA | | |
| | D56 CD4+ SARS-CoV-2 S2 | 0.79903597 | 0.81106632 | 0.03377386 | 0.00099194 | NA | |
| | D182 CD4+ SARS-CoV-2 S2 | 0.40095888 | 0.40896996 | 0.11921927 | 0.04722083 | 0.80650014 | NA |

**Supplementary table 2c:** Spearman's correlations and P values for proliferative CD8+ T cell response to SARS-CoV-2 S1

| | | D0 CD8+ SARS-CoV-2 S1 | D14 CD8+ SARS-CoV-2 S1 | D28 CD8+ SARS-CoV-2 S1 | D42 CD8+ SARS-CoV-2 S1 | D56 CD8+ SARS-CoV-2 S1 | D182 CD8+ SARS-CoV-2 S1 |
|---|---|---|---|---|---|---|---|
| Spearman's correlation | D0 CD8+ SARS-CoV-2 S1 | 1 | | | | | |
| | D14 CD8+ SARS-CoV-2 S1 | 0.65384615 | 1 | | | | |
| | D28 CD8+ SARS-CoV-2 S1 | 0.05454545 | 0.37979753 | 1 | | | |
| | D42 CD8+ SARS-CoV-2 S1 | 0.28181818 | 0.61986873 | 0.54022397 | 1 | | |
| | D56 CD8+ SARS-CoV-2 S1 | -0.07142857 | 0.36083863 | 0.46657929 | 0.3640855 | 1 | |
| | D182 CD8+ SARS-CoV-2 S1 | 0.00606061 | 0.29652642 | 0.16695652 | 0.11538462 | 0.29774436 | 1 |
| P value | D0 CD8+ SARS-CoV-2 S1 | NA | | | | | |
| | D14 CD8+ SARS-CoV-2 S1 | 0.01534852 | NA | | | | |
| | D28 CD8+ SARS-CoV-2 S1 | 0.88103618 | 0.03843964 | NA | | | |
| | D42 CD8+ SARS-CoV-2 S1 | 0.4011449 | 0.0002589 | 0.00067144 | NA | | |
| | D56 CD8+ SARS-CoV-2 S1 | 0.87904819 | 0.07638115 | 0.00814787 | 0.04405736 | NA | |
| | D182 CD8+ SARS-CoV-2 S1 | 0.98674291 | 0.18023947 | 0.43553811 | 0.58284984 | 0.20231999 | NA |

**Supplementary table 2d:** Spearman's correlations and P values for proliferative CD8+ T cell response to SARS-CoV-2 S2

| | | D0 CD8+ SARS-CoV-2 S2 | D14 CD8+ SARS-CoV-2 S2 | D28 CD8+ SARS-CoV-2 S2 | D42 CD8+ SARS-CoV-2 S2 | D56 CD8+ SARS-CoV-2 S2 | D182 CD8+ SARS-CoV-2 S2 |
|---|---|---|---|---|---|---|---|
| Spearman's correlation | D0 CD8+ SARS-CoV-2 S2 | 1 | | | | | |
| | D14 CD8+ SARS-CoV-2 S2 | 0.01176471 | 1 | | | | |
| | D28 CD8+ SARS-CoV-2 S2 | 0.03296703 | 0.67325617 | 1 | | | |
| | D42 CD8+ SARS-CoV-2 S2 | 0.08241758 | 0.82495765 | 0.50358974 | 1 | | |
| | D56 CD8+ SARS-CoV-2 S2 | -0.28333333 | 0.58668677 | 0.31586151 | 0.69379382 | 1 | |
| | D182 CD8+ SARS-CoV-2 S2 | 0.66666667 | 0.02197802 | -0.12745098 | -0.00294118 | -0.15011042 | 1 |
| P value | D0 CD8+ SARS-CoV-2 S2 | NA | | | | | |
| | D14 CD8+ SARS-CoV-2 S2 | 0.96550817 | NA | | | | |
| | D28 CD8+ SARS-CoV-2 S2 | 0.91485647 | 0.00059447 | NA | | | |
| | D42 CD8+ SARS-CoV-2 S2 | 0.78895093 | 2.32E-06 | 0.00872244 | NA | | |
| | D56 CD8+ SARS-CoV-2 S2 | 0.46003033 | 0.00654538 | 0.10154498 | 8.47E-05 | NA | |
| | D182 CD8+ SARS-CoV-2 S2 | 0.04986723 | 0.94318624 | 0.62593015 | 0.9913748 | 0.57896618 | NA |

**Supplementary table 3:** P values (by fisher test) for SARS-CoV-2 responders vs non-responders with CCC responses

| | | HKU1 clade 1 S1 | HKU1 clade 1 S2 | HKU1 clade 2 S1 | HKU1 clade 2 S2 | OC43 S1 | OC43 S2 | 299E S1 | 299E S2 | NL63 S1 | NL63 S2 |
|---|---|---|---|---|---|---|---|---|---|---|---|
| CD4+ | SARS-CoV-2 S1 | 0.342 | 0.143 | 0.342 | 0.033 | 0.342 | 0.036 | 0.242 | 0.064 | 0.481 | >0.999 |
| | SARS-CoV-2 S2 | 0.111 | >0.999 | 0.61 | 0.066 | 0.61 | 0.011 | 0.42 | 0.002 | >0.999 | >0.999 |
| CD8+ | SARS-CoV-2 S1 | 0.427 | 0.015 | 0.14 | 0.0004 | 0.242 | 0.584 | >0.9999 | 0.109 | >0.999 | 0.14 |
| | SARS-CoV-2 S2 | 0.083 | 0.0012 | 0.022 | 0.003 | 0.069 | 0.023 | 0.073 | 0.0007 | >0.999 | 0.206 |

**Supplementary table 4:** Spearman's correlations and P values for a panel of coronavirus antibody responses measured using MSD technology.

| | | D0 SARS-CoV-2 S | D0 SARS-CoV-1 S | D0 MERS-CoV S | D0 HKU-1 S | D0 OC43 S | D0 299E S | D0 NL63 S |
|---|---|---|---|---|---|---|---|---|
| Spearman's correlation | D0 SARS-CoV-2 S | 1 | | | | | | |
| | D0 SARS-CoV-1 S | 0.550054618 | 1 | | | | | |
| | D0 MERS-CoV S | 0.589027098 | 0.739252995 | 1 | | | | |
| | D0 HKU-1 S | 0.440184644 | 0.461451727 | 0.451162791 | 1 | | | |
| | D0 OC43 S | 0.040663871 | 0.332346723 | 0.45961945 | 0.520366455 | 1 | | |
| | D0 299E S | 0.159343176 | 0.256659619 | 0.146723044 | 0.40944327 | 0.469203665 | 1 | |
| | D0 NL63 S | 0.22854928 | 0.296969697 | 0.167159972 | 0.614235377 | 0.456095842 | 0.52572234 | 1 |
| P value | D0 SARS-CoV-2 S | NA | | | | | | |
| | D0 SARS-CoV-1 S | 0.000109783 | NA | | | | | |
| | D0 MERS-CoV S | 2.60E-05 | 9.98E-09 | NA | | | | |
| | D0 HKU-1 S | 0.002788824 | 0.001617085 | 0.002114079 | NA | | | |
| | D0 OC43 S | 0.79326225 | 0.027513953 | 0.00169716 | 0.000293719 | NA | | |
| | D0 299E S | 0.301531675 | 0.092611767 | 0.341914575 | 0.005782097 | 0.001314138 | NA | |
| | D0 NL63 S | 0.135643947 | 0.050279606 | 0.278122749 | 9.21E-06 | 0.001861083 | 0.000247587 | NA |

**Supplementary table 5:** T cell proliferation panel using Cell trace violet® (CTV) cell dye

| | Fluorochrome | Marker | Clone | Volume/50ul | Catalogue No | Company |
|---|---|---|---|---|---|---|
| CTV Proliferation Assay | | L/D NIR | | | L24976 | Invitrogen |
| | | CTV | | | C34557 | Invitrogen |
| | FITC | CD3 | UCHT1 | 1 | 300440 | Biolegend |
| | APC | CD4 | RPA-T4 | 0.25 | 300537 | Biolegend |
| | PE CY7 | CD8 | RPA-T8 | 0.25 | 301012 | Biolegend |

**Supplementary table 6:** Activation induced marker (AIM) panel

| | Fluorochrome | Marker | Clone | Volume/50ul | Catalogue No | Company |
|---|---|---|---|---|---|---|
| AIM assay | APC | CXCR3 | 1C6/CXCR3 | 5 | 550967 | BD |
| | BB515 | CXCR5 | RF8B2 | 1 | 564624 | BD |
| | APC CY7 | CCR6 | G034E3 | 2 | 353432 | Biolegend |
| | BV421 | PD1 | EH12.2H7 | 1 | 329920 | Biolegend |
| | BV510 | CD19 | HBI19 | 0.25 | 302242 | Biolegend |
| | BV510 | CD14 | M5E2 | 0.25 | 301842 | Biolegend |
| | PE CF 594 | CD137 | 4B4-1 | 1 | 309826 | Biolegend |
| | BV650 | CD69 | FN50 | 0.5 | 310934 | Biolegend |
| | BV570 | CD4 | SK3 | 1 | 300534 | Biolegend |
| | PERCP EFLUOR 710 | CD39 | eBioA1 | 1.5 | 46-0399-42 | Invitrogen |
| | PE | CD134 (OX40) | L106 | 2 | 340420 | BD |
| | | L/D AQUA | | | L34966 | Invitrogen |
| | PECY7 | CD25 | 2A3 | 1 | 335824 | BD |
| | APCR700 | CD8 | RPA-T8 | 0.25 | 565165 | BD |
| | BV605 | CD3 | UCHT1 | 0.5 | 300460 | Biolegend |

**Supplementary table 7:** Ex vivo activation panel

| | Fluorochrome | Marker | Clone | Volume/50ul | Catalogue No | Company |
|---|---|---|---|---|---|---|
| Ex vivo Activation Panel | L/D AQUA | Viability discriminator | | 1 | | |
| | BV510 | CD14 | M5E2 | 0.25 | 302242 | Biolegend |
| | BV510 | CD19 | HIB19 | 0.25 | 301842 | Biolegend |
| | BV6711 | CD3 | UCHT1 | 1 | 300464 | Biolegend |
| | BV570 | CD4 | RPA-T4 | 1 | 300534 | Biolegend |
| | BB515 | CXCR5 | RF8B2 | 1 | 564624 | BD Bioscience |
| | APC CY7 | HLA-DR | L243 | 1.25 | 307617 | Biolegend |
| | BB700 | CCR6 | 11A9 | 1 | 566478 | BD Bioscience |
| | APCR700 | CD8 | RPA-T8 | 1 | 565165 | BD Bioscience |
| | PE TxR | CD38 | HIT2 | 0.5 | 562288 | BD Bioscience |
| | APC | CXCR3 | IC6/CXCR3 | 10 | 550967 | BD Bioscience |

**Supplementary table 8:** Ex vivo exhaustion panel

| | Fluorochrome | Marker | Clone | Volume/50ul | Catalogue No | Company |
|---|---|---|---|---|---|---|
| T cell exhaustion Panel | L/D AQUA | Viability discriminator | | | | |
| | BV510 | CD14 | M5E2 | 0.25 | 302242 | Biolegend |
| | BV510 | CD19 | HIB19 | 0.25 | 301842 | Biolegend |
| | BV605 | CD3 | UCHT1 | 0.5 | 300460 | Biolegend |
| | BV570 | CD4 | RPA-T4 | 1 | 300534 | Biolegend |
| | BV421 | PD1 | EH12.2H7 | 1 | 329920 | Biolegend |
| | APCR700 | CD8 | RPA-T8 | 0.5 | 565165 | BD Bioscience |
| | PE TXR | EOMES | WD1928 | 1.25 | 61-4877-42 | Invitrogen |
| | FITC | TBET | 4B10 | 1.25 | 644812 | Biolegend |
